# Trends in Prescription Fills for Blood Pressure-Elevating and Antihypertensive Medications, 2017–2023

**DOI:** 10.1101/2024.11.14.24317362

**Authors:** Ashutosh Kumar, Nicole L. Therrien, John I. Ogwuegbu, Jun Soo Lee, Hilary K. Wall, John M. Flack, Sandra L. Jackson

**Author notes:** Corresponding author: Sandra L. Jackson; Chamblee 107, 4770 Buford Highway NE, Atlanta, GA 30341, USA. Financial Disclosures: None. Disclaimer: The findings and conclusions in this study are those of the authors and do not necessarily represent the official position of the Centers for Disease Control and Prevention (CDC).

## Abstract

**Background:** Many medications can have blood pressure (BP)-elevating effects, which might negatively impact BP control among persons with hypertension. This study examines trends in prescription fills for BP-elevating and antihypertensive medications, individually and concurrently, among US individuals.

**Methods:** Quarterly trends of concurrent and individual fills for BP-elevating and antihypertensive medications were reported using the national sample from IQVIA’s Total Patient Tracker database, covering 94% of all retail prescription fills in the US. We identified 1,387 products containing BP-elevating medications and 240 products containing antihypertensive medications. Percentage change from Q1/2017 and average quarterly percent change from Joinpoint regression were used to present trends overall and by sex and age group (0–17, 18–44, 45–64, 65–74, and ≥75 years).

**Results:** During 2017–2023, fills remained stable for BP-elevating medications alone and increased for antihypertensive medication alone (9.5% increase; from 10.1% to 11.0%; p<0.001). Concurrent fills for antihypertensive and BP-elevating medications increased by 15.9% (from 5.4% to 6.2%; p<0.001). Prescription fills for BP-elevating medications were higher among adult women compared to men; among women aged 18 to 44 years, this was primarily due to the use of combined oral contraceptives. In Q4/2023, prescription fills for BP-elevating medications were most common among those aged 65–74 years (females=30.7% vs males = 20.4%).

**Conclusions:** These results provide the first national trends in concurrent prescription fills for BP-elevating and antihypertensive medications, indicating an increasing trend. Our findings might inform clinicians in their decision-making regarding medication selection for patients with hypertension.

## INTRODUCTION

Hypertension, or high blood pressure (BP), is one of the major modifiable risk factors for cardiovascular disease (CVD) including heart disease and stroke,^1^ which are among the leading causes of death in the United States (US).^2,3^ Hypertension, is a chronic medical condition that affects a significant portion of the US population.^4^ According to 2017–20 estimates, nearly 48.1% of US adults have hypertension, of which 77.5% have uncontrolled hypertension (BP ≥ 130/80 mmHg).^4^ Hypertension control is critically important as it is significantly associated with lower risks of premature heart disease mortality^5,6^ and major acute cardiovascular events including myocardial infarction and stroke.^7,8^

One of the less investigated barriers in BP control is the use of medications that can raise BP levels.^9,10^ According to the 2017 American College of Cardiology and American Heart Association Guideline,^10^ certain prescription medications, such as amphetamines, antidepressants, non-steroidal anti-inflammatory drugs (NSAIDs), immunosuppressants, and combined oral contraceptives can raise blood pressure; use of these medications might lead to hypertension or worsening BP control among those with hypertension.^9–11^ Concurrent use of BP-elevating medication might attenuate antihypertensive effects of antihypertensive therapy or might also lead to a change in BP from drug interactions.^12^

Prior studies have documented antihypertensive medication trends among the US population;^13–16^ however, there is scarce literature on prescription fills for BP-elevating medications. One study, using the National Health and Nutrition Examination Survey (NHANES) pooled data from 2009 to 2018, found that approximately 18% of US adults with hypertension took medications that might cause elevated blood pressure.^9^ Antidepressants, NSAIDs, and estrogen-containing medications were the most commonly reported classes, and the use of these medications was associated with greater odds of hypertension among adults not concurrently taking antihypertensive medications. Understanding the prescription trends for concurrent BP-elevating and antihypertensive medications can provide insights into the strategies needed to improve hypertension management and control.

The primary objective of this study is to present quarterly trends of prescription fills for BP-elevating and antihypertensive medications, individually and concurrently, among US individuals from 2017 to 2023. Additionally, we present trends by sex and age groups for BP-elevating and antihypertensive medications.

## METHOD

### Data

We used IQVIA’s Total Patient Tracker (TPT) database to obtain the deduplicated counts of individuals with prescription fills across the US in each quarter from Q1/2017 to Q4/2023. Covering 94% of outpatient retail prescription fills in the US, the TPT database gathers prescription information directly from sources including payers and retail pharmacies such as pharmacy chains, independent pharmacies, grocery stores, and large retail outlets. The TPT database provides nationally projected deduplicated counts of individuals with prescription fills along with 95% confidence intervals. Consistent with previous studies using TPT data, prescriptions from veterinarians, dentists, and naturopaths were excluded.^17^

We identified 240 products containing antihypertensive medications (referred to as antihypertensive medications) (Table S1).^15^ For BP-elevating medications, a list of 1,387 products containing potentially BP-elevating medications (referred to as BP-elevating medications) was generated from a search of 150 generic drug names across 11 therapeutic classes (Table S2). Included generic drug names and therapeutic classes were guided by internal authors’, including pharmacists’, consensus, and were based on medications with potential BP-elevating effects, mentioned in the hypertension clinical practice guideline^10^ and a thorough review of the existing literature.^9,12,18–37^

The TPT database provided the overall counts of unique individuals for each quarter in three mutually exclusive groups: 1) prescription fills for BP-elevating medication alone, 2) prescription fills for antihypertensive medication alone, and 3) concurrent prescription fills for BP-elevating medication and antihypertensive medication. Concurrent prescriptions were defined if prescriptions for any of the BP-elevating and antihypertensive medications were filled in the same quarter. Furthermore, we obtained the counts of unique individuals by sex and age group for each quarter who had individual or concurrent prescription fills for any antihypertensive medication or any BP-elevating medication (i.e., non-mutually exclusive categories). To report results by sex and age group, we presented findings for a total of 10 subgroups: among males and females in 5 age ranges (0–17, 18–44, 45–64, 65–74, and ≥75 years). All data were deidentified and no institutional board approval was required.

### Statistical analysis

We reported deduplicated count of individuals with concurrent or individual prescription fills for BP-elevating and antihypertensive medications for each quarter from Q1/2017 to Q4/2023. We used annual US population totals from the CDC WONDER dataset provided by National Center for Health Statistics (NCHS)^38^ for each group to report the age-specific prevalence of prescription fills in percentages, absolute (non-regression based) percent changes compared to Q1/2017, and quarterly trends, overall and stratified by sex and age group. We used the NCHS 2022 population data for population denominators for our data in both 2022 and 2023, as NCHS population data for 2023 was not yet available. Data extraction and analysis was conducted using Microsoft Excel and Stata, version 17.0 (StataCorp).

Joinpoint regression (JoinPoint 5.1.0, National Cancer Institute, Bethesda, MD) identified statistically significant trends in population percentage from Q1/2017 to Q4/2023.^39^ For Joinpoint regression, a permutation-based approach was adopted for all groups, and the log model with correlated errors was selected.^40,41^ Estimates from Joinpoint regression for the population percentage with prescription fills were reported as the average quarterly percent change (AQPC) for the period Q1/2017–Q4/2023. We excluded the year 2020 in estimating trends and statistical significance from the Joinpoint regression.^42^ Estimates and statistical significance were similar to the Joinpoint regression model with the full sample (Table S3).

Furthermore, we presented absolute change for the top 20 medications within antihypertensive and BP-elevating medication groups that experienced largest increase in number of individuals with prescription fills by sex and age group from Q1/2017 to Q4/2023 (Tables S4 and S5). We also presented the list of top 20 medications with highest number of fills in Q4/2023 within antihypertensive and BP-elevating medication (Tables S6 and S7). We reported population percentage and Joinpoint results with 95% confidence intervals, and statistical significance was determined by a p-value <0.05. We followed the Strengthening the Reporting of Observational Studies in Epidemiology (STROBE) reporting guidelines.

## RESULTS

### Overall results for concurrent and individual prescription fills

Prescription fills for BP-elevating medications alone remained stable from 12.5% (95% CI = 12.4% to 12.6%) (40.8 million) in Q1/2017 to 12.7% (95% CI = 12.6% to 12.8%) (42.3 million) in Q4/2023; however, prescription fills for antihypertensive medications alone and concurrent prescription fills for BP-elevating and antihypertensive medications increased during Q1/2017 to Q4/2023 (Figure 1, Tables 1 and 2). The percentage of individuals with prescription fills for antihypertensive medication alone increased from 10.1% (95% CI = 10.0% to 10.2%) (32.8 million) in Q1/2017 to 11.0% (95% CI = 10.9% to 11.1%) (36.8 million) in Q4/2023 (9.5% increase; p<0.001). Concurrent prescription fills for BP-elevating and antihypertensive medications increased from 5.4% (95% CI = 5.3% to 5.4%) (17.4 million) in Q1/2017 to 6.2% (95% CI = 6.1% to 6.3%) (20.7 million) in Q4/2023 (15.9% increase; p<0.001).

**Figure 1:**
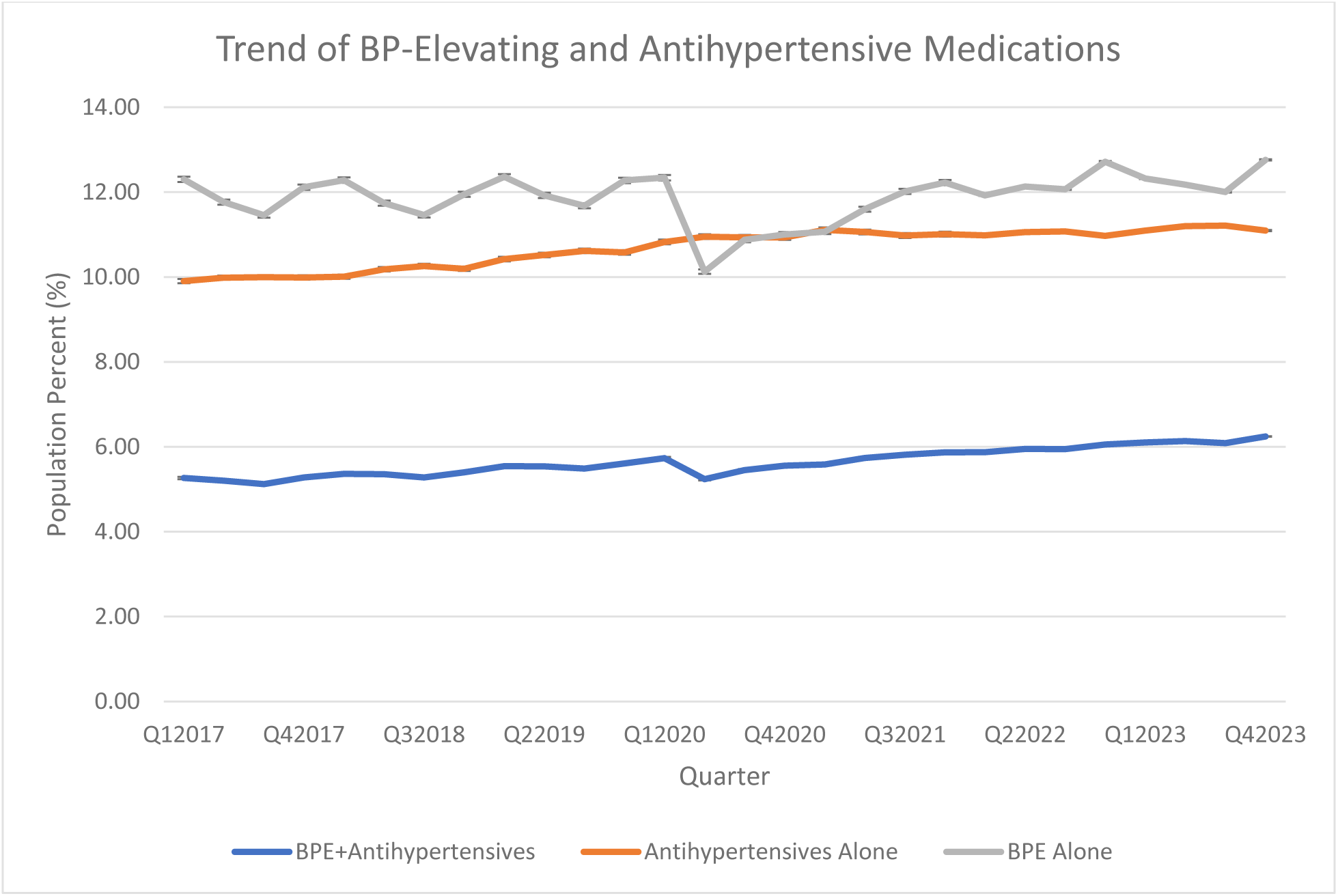
Population percent with unique number of individuals with prescription fills for any BP-elevating medication alone, any antihypertensive medication alone, and concurrent prescription fills for any BP-elevating plus any antihypertensive medication from IQVIA’s TPT Q1, 2017–Q4, 2023^a^ ^a^ This figure shows the quarterly trend of US population percentage for unique number of individuals with prescription fills for BP-elevating medications only, antihypertensive medications only, and concurrent prescription fills for any antihypertensive and any BP-elevating medications using IQVIA’s Total Patient Tracker database, Q1/2017 to Q4/2023. Total US population was used to obtain population percentage for all quarters. Drug prescriptions for unspecified gender and age groups were excluded. The total aggregate numbers are slightly lower than the sum of numbers from subgroups as the deduplication process removes multiple entries for the same person appearing in different age bands while removing all conflicting/missing reports of sex and age groups.

**Figure 2:**
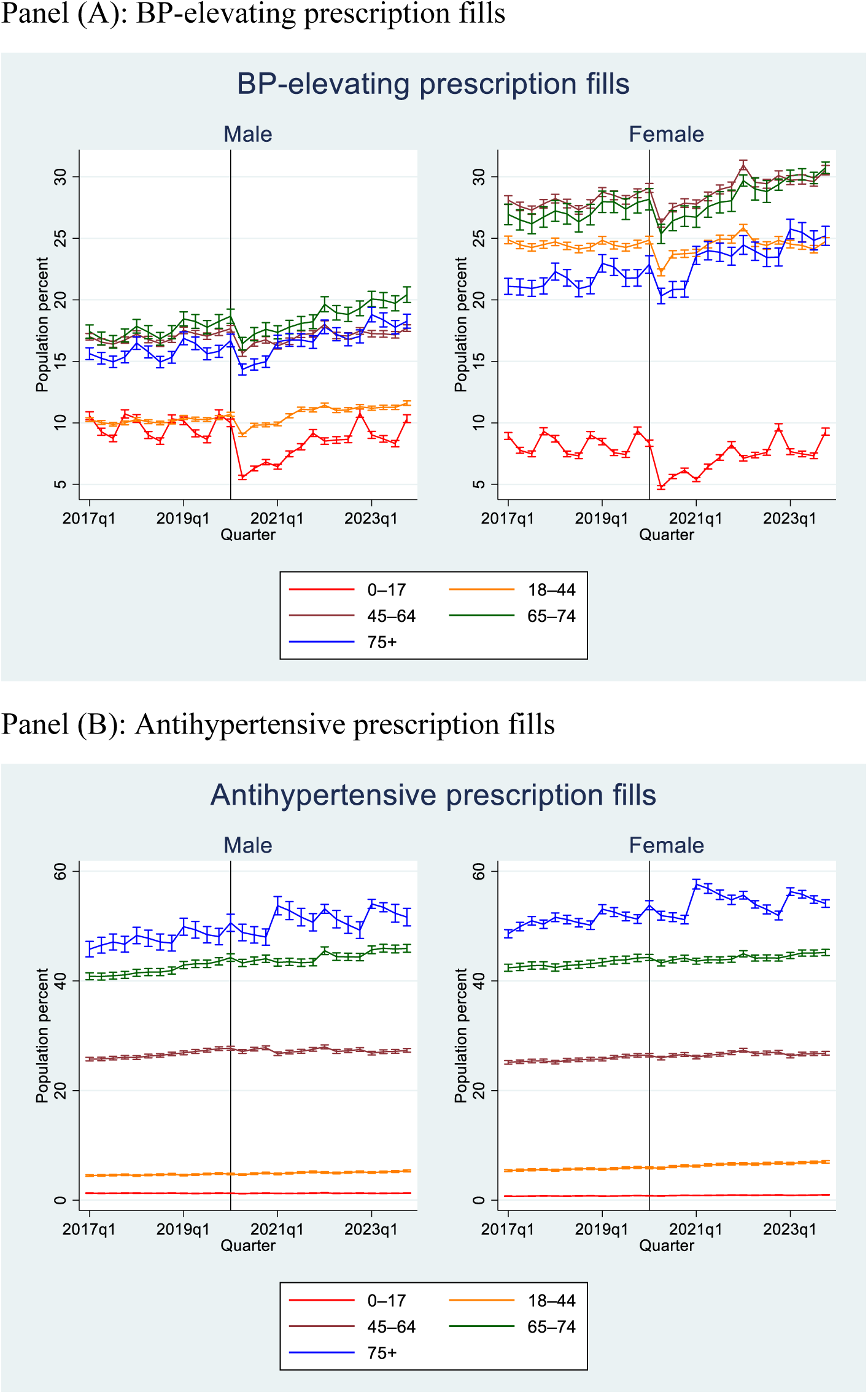
Projected percentage of population with prescription fills for BP-elevating and antihypertensive medication from outpatient retail pharmacies, stratified by sex and age group from IQVIA’s TPT Q1, 2017—Q4, 2023^a^ ^a^ This figure presents population percentage of individuals with prescription fills for BP-elevating (panel A) and antihypertensive medication (panel B) from Q1/2017 to Q4/2023. Annual US population totals from the CDC WONDER dataset provided by National Center for Health Statistics (NCHS) for each group were used to report the age-specific prevalence of prescription fills in percentages. Since population denominator for year 2023 was not available, hence, population total of year 2022 was used as population denominator for both years 2022 and 2023. IQVIA’s TPT portal was used to obtain deduplicated counts of individuals with prescription fill with 95% confidence interval.

**Table 1:** Number of unique individuals and population percent with prescription fills for any BP- elevating medication alone, any antihypertensive medication alone, and concurrent prescription fills for any BP-elevating plus any antihypertensive medication in Q1/2017 and Q4/2023 from IQVIA’s TPT Q1, 2017–Q4, 2023^a^

|  | Q1/2017 |  |  |  | Q4/2023 |  |  |  |  |
| --- | --- | --- | --- | --- | --- | --- | --- | --- | --- |
| Group | Count | Pop. percent | 95% lower CI | 95% upper CI | Count | Pop. percent | 95% lower CI | 95% upper CI | Percent change from Q1/2017 to Q4/2023<sup>b</sup> |
| **Overall** |  |  |  |  |  |  |  |  |  |
| BPE Alone | 40775287 | 12.5 | 12.4 | 12.6 | 42293817 | 12.7 | 12.6 | 12.8 | 1.4 |
| Anti-HTN Alone | 32823564 | 10.1 | 10.0 | 10.2 | 36773683 | 11.0 | 10.9 | 11.1 | 9.5* |
| Anti-HTN + BPE Concurrent | 17449846 | 5.4 | 5.3 | 5.4 | 20694112 | 6.2 | 6.1 | 6.3 | 15.9* |
| **Female, Prescription fills for BP-elevating medications (alone or concurrent)** |  |  |  |  |  |  |  |  |  |
| 0–17 | 3218663 | 8.9 | 8.7 | 9.2 | 3284969 | 9.3 | 9.0 | 9.6 | 4.0 |
| 18–44 | 14371855 | 24.9 | 24.6 | 25.2 | 14648654 | 24.7 | 24.4 | 25.1 | -0.5 |
| 45–64 | 12144460 | 28.1 | 27.8 | 28.5 | 12694235 | 30.5 | 30.2 | 30.9 | 8.7* |
| 65–74 | 4258200 | 26.9 | 26.1 | 27.8 | 5495503 | 30.7 | 30.3 | 31.2 | 14.1* |
| ≥75 | 2634337 | 21.1 | 20.4 | 21.7 | 3524894 | 25.2 | 24.4 | 26.0 | 19.5* |
| **Male, Prescription fills for BP-elevating medications (alone or concurrent)** |  |  |  |  |  |  |  |  |  |
| 0–17 | 3978019 | 10.6 | 10.2 | 10.9 | 3833869 | 10.3 | 10.0 | 10.7 | -2.2 |
| 18–44 | 6059425 | 10.3 | 10.1 | 10.4 | 7117764 | 11.6 | 11.4 | 11.8 | 13.1 |
| 45–64 | 6999287 | 17.0 | 16.7 | 17.3 | 7250265 | 17.7 | 17.4 | 18.0 | 4.1* |
| 65–74 | 2417488 | 17.4 | 16.9 | 18.0 | 3248851 | 20.4 | 19.8 | 21.1 | 17.2* |
| ≥75 | 1357037 | 15.6 | 15.1 | 16.1 | 1830013 | 18.3 | 17.7 | 18.8 | 16.9* |
| **Female, Prescription fills for antihypertensive medications (alone or concurrent)** |  |  |  |  |  |  |  |  |  |
| 0–17 | 262530 | 0.7 | 0.7 | 0.8 | 346233 | 1.0 | 0.9 | 1.0 | 34.3* |
| 18–44 | 3106753 | 5.4 | 5.2 | 5.5 | 4133050 | 7.0 | 6.8 | 7.2 | 29.8* |
| 45–64 | 10862267 | 25.1 | 24.8 | 25.5 | 11142799 | 26.8 | 26.5 | 27.1 | 6.6* |
| 65–74 | 6702170 | 42.4 | 41.7 | 43.1 | 8078775 | 45.2 | 44.6 | 45.8 | 6.6* |
| ≥75 | 6069932 | 48.6 | 47.9 | 49.4 | 7569161 | 54.1 | 53.4 | 54.8 | 11.3 |
| **Male, Prescription fills for antihypertensive medications (alone or concurrent)** |  |  |  |  |  |  |  |  |  |
| 0–17 | 483457 | 1.3 | 1.2 | 1.3 | 479766 | 1.3 | 1.3 | 1.3 | 0.7 |
| 18–44 | 2652720 | 4.5 | 4.4 | 4.6 | 3267689 | 5.3 | 5.2 | 5.5 | 18.6* |
| 45–64 | 10594263 | 25.7 | 25.4 | 26.1 | 11196741 | 27.3 | 27.0 | 27.7 | 6.2* |
| 65–74 | 5667990 | 40.8 | 40.2 | 41.5 | 7308480 | 45.9 | 45.2 | 46.6 | 12.5* |
| ≥75 | 3978876 | 45.8 | 44.4 | 47.2 | 5173761 | 51.6 | 50.0 | 53.2 | 12.8* |

Abbreviations: BPE, blood pressure-elevating; Anti-HTN, antihypertensive; CI, confidence interval; Pop, population
<sup>a</sup>This table presents the prescription fills for BP-elevating and antihypertensive medications for Q1/2017 and Q4/2023 from IQVIA's Total Patient Tracker database.
<sup>b</sup>The percent change is obtained by comparing the population percent from Q1/2017 to Q4/2023. Percent change with statistical significance of p-value <0.05 from Joinpoint regression are highlighted with asterisk (\*) sign.

**Table 2:** Joinpoint regression results along with absolute change in Q4/2023 in comparison to Q1/2017 for concurrent and individual prescription fills for blood pressure (BP)-elevating and antihypertensive medications overall and by sex and age groups

| Group | Number of join points | Slope change quarter | AQPC<sup>a</sup> | 95% lower CI | 95% upper CI | AQPC<sup>a</sup> P-value | Percent change from Q1, 2017 to Q4, 2023 |
| --- | --- | --- | --- | --- | --- | --- | --- |
| **Overall** |  |  |  |  |  |  |  |
| BPE alone | 0 | NA | 0.02 | -0.11 | 0.2 | 0.7 | 1.4 |
| Anti-HTN alone | 1 | Q17 | 0.4* | 0.3 | 0.4 | <0.001 | 9.5* |
| Anti-HTN + BPE concurrent | 0 | NA | 0.6* | 0.5 | 0.6 | <0.001 | 15.9* |
| **Female, Prescription fills for BP-elevating medications (alone or concurrent)** |  |  |  |  |  |  |  |
| 0 to 17 | 0 | NA | -0.3 | -0.8 | 0.3 | 0.3 | 4.0 |
| 18 to 44 | 2 | Q17, Q20 | -0.1 | -0.6 | 0.3 | 0.8 | -0.5 |
| 45 to 64 | 0 | NA | 0.4* | 0.3 | 0.4 | <0.001 | 8.7* |
| 65 to 74 | 0 | NA | 0.5* | 0.5 | 0.6 | <0.001 | 14.1* |
| ≥75 | 0 | NA | 0.7* | 0.6 | 0.8 | <0.001 | 19.5* |
| **Male, Prescription fills for BP-elevating medications (alone or concurrent)** |  |  |  |  |  |  |  |
| 0 to 17 | 0 | NA | -0.4 | -0.9 | 0.1 | 0.1 | -2.2 |
| 18 to 44 | 2 | Q17, Q20 | 0.5 | -0.1 | 1.0 | 0.1 | 13.1 |
| 45 to 64 | 0 | NA | 0.1* | 0.1 | 0.2 | 0.004 | 4.1* |
| 65 to 74 | 0 | NA | 0.6* | 0.5 | 0.7 | <0.001 | 17.2* |
| ≥75 | 0 | NA | 0.7* | 0.5 | 0.8 | <0.001 | 16.9* |
| **Female, Prescription fills for antihypertensive medications (alone or concurrent)** |  |  |  |  |  |  |  |
| 0 to 17 | 0 | NA | 1.1* | 0.9 | 1.2 | <0.001 | 34.3* |
| 18 to 44 | 2 | Q9, Q20 | 0.9* | 0.8 | 1.0 | <0.001 | 29.8* |
| 45 to 64 | 1 | Q21 | 0.2* | 0.1 | 0.3 | <0.002 | 6.6* |
| 65 to 74 | 0 | NA | 0.2* | 0.2 | 0.2 | <0.001 | 6.6* |
| ≥75 | 4 | Q11, Q18, Q23, Q26 | 0.3 | -0.4 | 1.0 | 0.4 | 11.3 |
| **Male, Prescription fills for antihypertensive medications (alone or concurrent)** |  |  |  |  |  |  |  |
| 0 to 17 | 0 | NA | 0.0 | -0.1 | 0.1 | 1.0 | 0.7 |
| 18 to 44 | 0 | NA | 0.6* | 0.5 | 0.6 | <0.001 | 18.6* |
| 45 to 64 | 1 | Q12 | 0.2* | 0.1 | 0.3 | <0.001 | 6.2* |
| 65 to 74 | 0 | NA | 0.4* | 0.4 | 0.5 | <0.001 | 12.5* |
| ≥75 | 0 | NA | 0.6* | 0.4 | 0.7 | <0.001 | 12.8* |

Abbreviations: Anti-HTN, antihypertensive; AQPC, average quarterly percent change; BPE, blood pressure-elevating, NA, not applicable
<sup>a</sup> AQPC values are obtained from Joinpoint regression after excluding the data for year 2020. P-values <0.05 were considered statistically significant and are highlighted by asterisk (\*) sign.

### Prescription fills for BP-elevating medications overall and by sex and age groups

Prescription fills for BP-elevating medications were higher among females in comparison to males for all age groups except those aged 0–17 years (Figure 1, Tables 1 and 2). For example, in Q4/2023, among women of reproductive age (18–44 years), 24.7% (95%CI= 24.4% to 25.1%) had fills for BP-elevating medications, compared to only 11.6% (95%CI= 11.4% to 11.8%) of men of the same age group. Similarly, prescription fills for BP-elevating medications in Q4/2023 were higher among females in comparison to males for other adult age groups — 30.5% (95%CI= 30.2% to 30.9%) vs 17.7% (95%CI= 17.4% to 18.0%) among 45–64 years, 30.7% (95%CI= 30.3% to 31.2%) vs 20.4% (95%CI= 19.8% to 21.1%) among 65–74 years, and 25.2% (95%CI= 24.4% to 26.0%) versus 18.3% (95%CI= 17.7% to 18.8%) among ≥75 years.

From Q1/2017 to Q4/2023, prescription fills for BP-elevating medications increased among males and females aged 45 years or above. Among females, the sharpest increase in prescription fills for BP-elevating medications occurred among those aged ≥75 years (19.5% increase; from 21.1% to 25.2%; p<0.001), followed by those aged 65–74 years (14.1% increase, from 26.9% to 30.7%, p<0.001), and 45–64 years (8.7% increase, from 28.1% to 30.5%, p<0.001). Among males, the sharpest increase in prescription fills for BP-elevating medications occurred among those aged 65–74 years (17.2% increase; from 17.4% to 20.4%; p<0.001), followed by those aged ≥75 years (16.9% increase; from 15.6% to 18.3%; p<0.001), and 45–64 years (4.1% increase; from 28.1% to 30.5%; p=0.004).

Among BP-elevating medications with largest increase in prescription fills from Q1/2017 to Q4/2023 for females, most common therapeutic classes were stimulants, atypical antipsychotics, antidepressants, estrogen-containing medications, and corticosteroids (Tables S4 and S6). Among males, most common therapeutic classes were stimulants, atypical antipsychotics, corticosteroids, and NSAIDs.

### Prescription fills for antihypertensive medications overall and by sex and age groups

Prescription fills for antihypertensive medications were higher for older age groups and were similar by age group for both sexes. For example, in Q4/2023 prescription fills for antihypertensive medications were highest among those aged ≥75 years (females = 54.1% vs males = 51.6%) followed by those aged 65–74 years (females = 45.2% vs males = 45.9%), 45– 64 years (females = 26.8% vs males = 27.3%), 18–44 years (females = 7.0% vs males = 5.3%), and 0–17 years (females = 1.0% vs males = 1.3%). However, there was some variation in trends in prescription fills for antihypertensive medication by sex and age group over time. From Q1/2017 to Q4/2023, prescription fills for antihypertensive medications increased among all sex and age groups, except among females aged ≥75 years and among males aged 0–17 years.

Among females, the sharpest increase in prescription fills for antihypertensive medications occurred among those aged 0–17 years (34.3% increase; from 0.7% to 1.0%; p<0.001), followed by those aged 18–44 years (29.8% increase; from 5.4% to 7.0%; p<0.001), 45–64 years (6.6% increase; from 25.1% to 26.8%; p<0.002), and 65–74 years (6.6% increase; from 42.4% to 45.2%; p<0.001). Among males, the sharpest increase in prescription fills for antihypertensive medications occurred among those aged 18–44 years (18.6% increase; from 4.5% to 5.3%; p<0.001), followed by those aged ≥75 years (12.8% increase; from 45.8% to 51.6%;p<0.001), 65–74 years (12.5% increase; from 40.8% to 45.9%; p<0.001), and 45–64 years (6.2% increase; from 25.7% to 27.3%; p<0.001).

Among antihypertensive medications with the largest increases in prescription fills from Q1/2017 to Q4/2023, most common therapeutic classes among females and males were angiotensin receptor blockers, beta blockers, and calcium channel blockers (Tables S5 and S7).

## DISCUSSION

This is the first study to present national trends of BP-elevating and antihypertensive medications, individually and concurrently, in the US. We provided recent trends across 28 quarters of data, using comprehensive lists of BP-elevating and antihypertensive medications. There were several key findings. First, about one out of every twenty persons in the US filled concurrent prescriptions for BP-elevating and antihypertensive medications, and concurrent fills increased by 15.9% during the study period. Second, nearly one in five persons in the US filled a prescription for a BP-elevating medication, either alone or concurrently with an antihypertensive medication, and fills for BP-elevating medications were higher among females than males for all adult age groups. Third, there was a significant rise in antihypertensive prescription fills among females under 45 years old and males aged 18–44 years.

In a recent study, Vitarello et al. reported that 14.9% of US adults and 18.5% of US adults with hypertension used medications that can elevate blood pressure during 2009–2018.^9^ We observed that the prevalence of prescription fills for BP-elevating medications was 17.9% (12.5% individually and 5.4% concurrently) in Q1/2017, which rose to 18.9% (12.7% individually and 6.1% concurrently) in Q4/2023. Our prevalence estimates for BP-elevating medications were higher than Vitarello et al. This difference could be due to our inclusion of a wider range of BP-elevating medications or our use of more recent data. We reported 15.9% increase in concurrent prescription fills for BP-elevating and antihypertensive medications during 2017–2023; such fills could hinder BP control for persons with hypertension. Recent estimates from 2013–2014 to 2017–2018 indicate a decline in BP control from 53.8% to 43.7% among adults with hypertension and from 72.2% to 64.8% among adults taking antihypertensive medication.^8^ The growth in concurrent BP-elevating and antihypertensive medication use might be a contributing factor in these trends. While many BP-elevating medications are beneficial or even life-saving, for individual patients with hypertension, clinicians and healthcare teams can examine concurrent prescriptions, consider risks and benefits of BP-elevating medications, and consider alternatives for BP-elevating medications when appropriate.

For all adult age groups in Q4/2023, we observed a higher percentage of prescription fills for BP-elevating medications among women (approximately 25–31%) compared to men (11– 20%). The higher percentage of prescription fills for BP-elevating medications among females aged 18 to 44 years was primarily due to the use of estrogen-containing medications, specifically combined oral contraceptives. Estimates from 2017–2019 indicate that the most common contraceptive methods among females aged 15–49 years in the US included contraceptive pills (14.0%, although the study did not distinguish between types of oral contraceptives).^43^ For other age groups among females, the largest increases in prescription fills occurred for medication classes such as stimulants, atypical antipsychotics, antidepressants, and corticosteroids. Prior studies have shown that a higher percentage of women than men use potentially BP-elevating medications such as antidepressants,^44^ NSAIDs,^45,46^ stimulants,^47^ and antipsychotics.^48–50^ For example, a higher percentage of women (17.7%) used an antidepressant than men (8.4%) during 2015–2018.^44,51^ Higher usage of BP-elevating medication among women than men is concerning as it might negatively affect BP control in this population. According to recent findings on BP control among adults with hypertension, BP control declined significantly among women (from 56.3% in 2009–2012 to 47.9% in 2017–2020) in comparison to no significant change among men (from 50.4% in 2009–2012 to 49.0% in 2017–2020).^52^

During 2017–2023, prescription fills for antihypertensive medications increased over time across most sex and age groups. Although prescription fills for antihypertensive medications were higher among older age groups, consistent with their higher prevalence of hypertension,^4^ the most meaningful *increase* in prescription fills for antihypertensive medications was observed among younger age groups. Our estimates suggest an increasing trend of prescription fills for antihypertensive medications among females (29.8% increase) and males (18.6% increase) of reproductive age (18–44 years) from Q1/2017 to Q4/2023. Although, to our knowledge, there are no recent estimates of antihypertensive usage by age- and sex-specific groups for the US population overall, our results are consistent with recent estimates of antihypertensive usage among adults with hypertension. For example, Sekkarie et al.^16^ reported that antihypertensive usage among adults with hypertension increased for women (from 64.4% to 68.5%) and men (from 56.8% to 59.4%) during 2017–2021. Similar to our findings, Sekkarie et al. documented the highest increase in antihypertensives usage among persons aged 18–44 years (from 38.0% to 42.5%) during 2017–2021.^16^ Given the increasing percentage of reproductive age women taking antihypertensive medications, these women and their medical providers may need to be aware of the clinical guidance for the safe use of contraception among women with personal characteristics and medical conditions such as hypertension when providing contraceptive counseling.^53^

The findings of this study have important implications for public health and clinical practice. Establishing clinical guidance for managing concurrent BP-elevating and antihypertensive medication use could help mitigate adverse effects on BP control. For patients with hypertension, mitigation strategies could include alternative treatments when appropriate, or potential intensification of antihypertensive therapy when BP-elevating medication is needed and conflicting effects on BP are unavoidable. Clinical decision support tools that screen for medications with divergent effects on BP could help facilitate tailored medication regimens to support BP management. Lastly, pharmacotherapy research is needed to understand if certain antihypertensive medications are more effective at lowering BP when various classes of BP-elevating medications are being taken.

## Limitations

This study has limitations. First, although the TPT database covers 94% of outpatient retail prescriptions and provides nationally projected estimates, it excludes mail-order prescription fills and does not include cash purchases or over-the-counter medications. Second, concurrent fill data were not available by sex and age group from the TPT database. Thus, we were unable to examine concurrent prescription fills for BP-elevating and antihypertensive medications by sex and age groups. Third, dosage, duration, and frequency of administration information was unavailable, and we were unable to determine whether or how these medications were taken. For example, the concurrent prescription fills only included cases when the prescription fills for both BP-elevating and antihypertensive medications occurred in the same quarter; however, it could be possible that the medications were not taken at the same time. Additionally, some BP-elevating medications are typically taken on an as-needed basis while others are taken daily. We could not, for example, exclude BP-elevating medications taken at such low doses or for such short durations that they might not have meaningfully impacted BP. Fourth, we did not have data on other critical factors that could have potentially influenced prescription patterns, such as income, race, and region. Fifth, we were unable to identify the underlying conditions or diagnoses for which these medications were prescribed, so, for example, we could not restrict analyses to persons with hypertension. Sixth, while BP-elevating effects of some therapeutic classes are well-established, there is limited evidence documenting the prevalence, magnitude, and biological mechanism of BP elevation for some therapeutic classes of the included BP-elevating medications. Future studies can address the limitations of this study to provide a more nuanced understanding of concurrent and individual prescription fills for BP-elevating and antihypertensive medications.

## CONCLUSIONS

Hypertension is a modifiable cardiovascular disease risk factor affecting nearly half of US adults, and over half of adults with hypertension have uncontrolled BP. This study reported a rising national trend in concurrent prescription fills for BP-elevating and antihypertensive medications, and a higher use of BP-elevating medications among adult women compared to men. Concurrent use of BP-elevating and antihypertensive medications might hinder efforts to control BP among persons with hypertension. Evidence from this study could inform clinicians and patients as they consider medication options and hypertension management strategies.

## Data Availability

This study uses proprietary data from IQVIA. Request for data can be obtained from IQVIA.

## Acknowledgements

This study was reviewed by the Centers for Disease Control and Prevention and conducted consistent with applicable federal law and the Centers for Disease Control and Prevention policy (5 C.F.R. part 46, 21 C.F.R. part 56; 42 U.S.C. Sect. 241(d); 5 U. S.C. Sect. 552a; 44 U.S.C. Sect. 3501 et seq.).

## Online Only Supplement: Tables and Figures

**Table S1:**
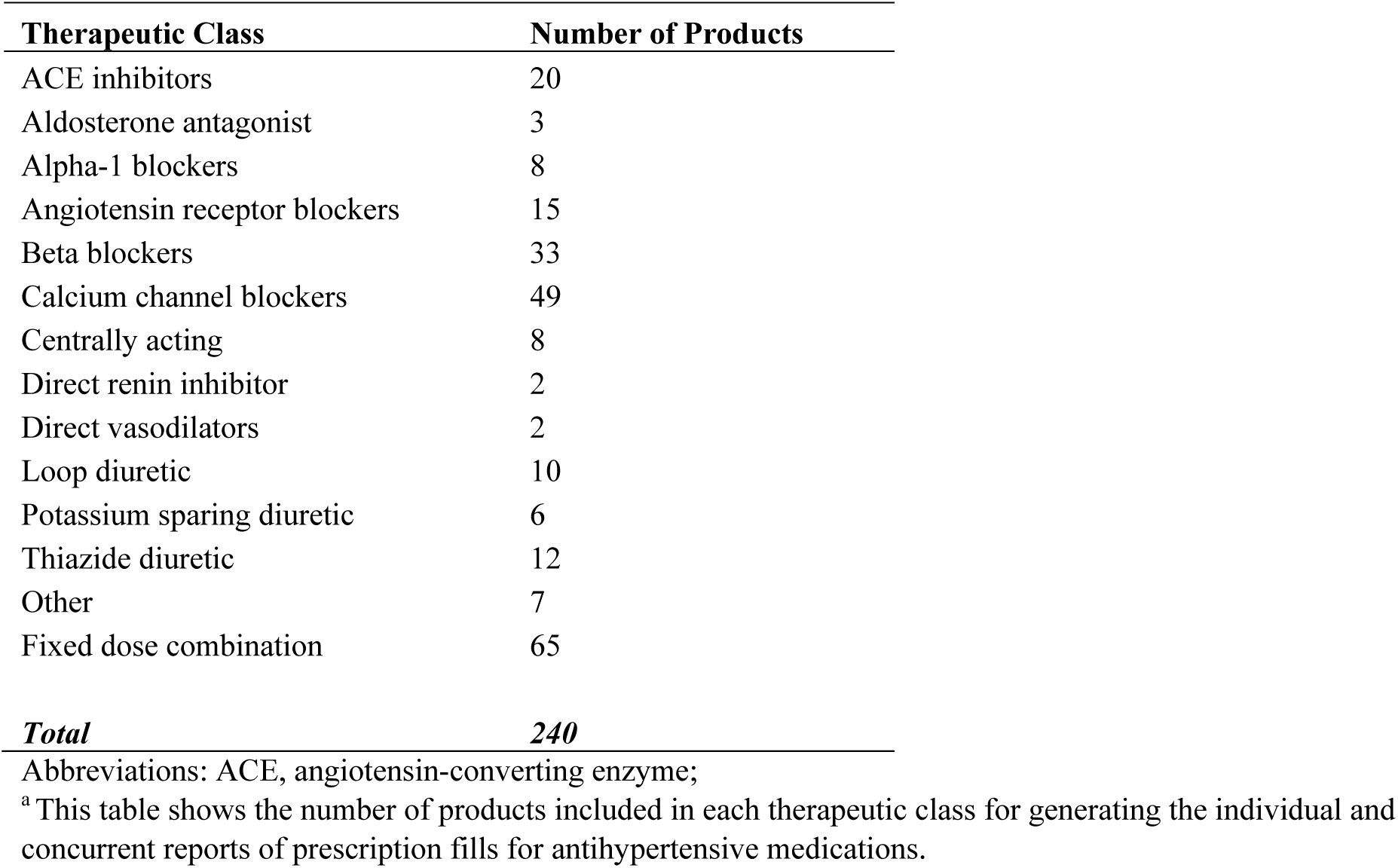
Therapeutic classes and corresponding number of products containing antihypertensive medications included in reporting the prescription fills overall and for sex and age groups from IQVIA’s Total Patient Tracker (TPT) database Q1, 2017—Q4, 2023^a^

| **Therapeutic Class** | **Number of Products** |
| --- | --- |
| ACE inhibitors | 20 |
| Aldosterone antagonist | 3 |
| Alpha-1 blockers | 8 |
| Angiotensin receptor blockers | 15 |
| Beta blockers | 33 |
| Calcium channel blockers | 49 |
| Centrally acting | 8 |
| Direct renin inhibitor | 2 |
| Direct vasodilators | 2 |
| Loop diuretic | 10 |
| Potassium sparing diuretic | 6 |
| Thiazide diuretic | 12 |
| Other | 7 |
| Fixed dose combination | 65 |
| ***Total*** | ***240*** |

Abbreviations: ACE, angiotensin-converting enzyme;
<sup>a</sup>This table shows the number of products included in each therapeutic class for generating the individual and concurrent reports of prescription fills for antihypertensive medications.

**Table S2:** Therapeutic classes and corresponding number of products containing blood pressure-elevating medications included in reporting the prescription fills overall and for sex and age groups from IQVIA’s Total Patient Tracker (TPT) database Q1, 2017—Q4, 2023^a,b^

| **Therapeutic Class** | **Number of Products** |
| --- | --- |
| Androgens | 30 |
| Antidepressants<sup>c</sup> | 59 |
| Antineoplastic Agent | 43 |
| Antiretrovirals | 72 |
| Atypical Antipsychotics | 51 |
| Corticosteroids | 66 |
| Decongestants | 529 |
| Estrogen-containing | 254 |
| Immunomodulators | 25 |
| NSAIDs | 196 |
| Stimulants | 62 |
| ***Total*** | ***1,387*** |

<sup>a</sup> This table shows the number of products included in each therapeutic class for generating the individual and concurrent reports of prescription fills for BP-elevating medications.
<sup>b</sup> Careful efforts were taken to remove topical products with limited systemic absorption. However, for BPE medication, the results were generated at the molecule level instead of the therapeutic class, and might include topical products with limited systemic absorption such as topical isotretinoin.
<sup>c</sup> The following antidepressant sub-classes were included in this analysis: Aminoketone, Monoamine Oxidase Inhibitors (MAOI), Serotonin-norepinephrine reuptake inhibitors (SNRI), Tricyclic antidepressants (TCA).

**Table S3:** Joinpoint regression estimates for full sample and for the sample excluding year 2020.

| Full sample |  |  | Excluding 2020 |  |  |
| --- | --- | --- | --- | --- | --- |
| Number of<br>Joinpoints | Mean square<br>errors | Average<br>quarterly percent<br>change (AQPC) | Number of<br>Joinpoints | Mean square<br>errors | Average<br>quarterly percent<br>change (AQPC) |
| ***Overall*** |  |  |  |  |  |
| 2 | 69.2938 | 0.1803 | 0 | 41.1271 | 0.0254 |
| 2 | 1.626 | 0.3286* | 1 | 1.9441 | 0.3592* |
| 2 | 13.8006 | 0.6306* | 0 | 5.9135 | 0.5609* |
| ***Prescription fills for BP-elevating medications, Female (alone or concurrent)*** |  |  |  |  |  |
| 2 | 72.4177 | 0.3156 | 0 | 75.8871 | -0.2689 |
| 2 | 14.2413 | -0.0876 | 2 | 5.0196 | -0.0574 |
| 3 | 11.1429 | 0.306 | 0 | 10.5366 | 0.3450* |
| 3 | 2.2435 | 0.5354 | 0 | 3.0496 | 0.5409* |
| 3 | 5.1013 | 0.6711 | 0 | 2.8621 | 0.7054* |
| ***Prescription fills for BP-elevating medications, Male (alone or concurrent)*** |  |  |  |  |  |
| 2 | 68.5309 | 0.21 | 0 | 67.9991 | -0.4138 |
| 3 | 14.0041 | 0.498 | 2 | 5.9916 | 0.4456 |
| 0 | 17.7451 | 0.1358* | 0 | 9.0166 | 0.1424* |
| 2 | 3.3122 | 0.7382 | 0 | 3.9473 | 0.5948* |
| 1 | 9.369 | 0.6485* | 0 | 4.5397 | 0.6455* |
| ***Prescription fills for antihypertensive medications, Female (alone or concurrent)*** |  |  |  |  |  |
| 0 | 3.4538 | 1.09* | 0 | 2.841 | 1.0792* |
| 2 | 0.6147 | 0.9212* | 2 | 0.5374 | 0.9089* |
| 1 | 2.456 | 0.2134* | 1 | 2.0337 | 0.2150* |
| 0 | 1.7539 | 0.2049* | 0 | 1.3062 | 0.2079* |
| 3 | 13.6629 | 0.3782 | 4 | 8.3083 | 0.3009 |
| ***Prescription fills for antihypertensive medications, Male (alone or concurrent)*** |  |  |  |  |  |
| 0 | 1.6419 | -0.0102 | 0 | 1.5285 | 0.0001 |
| 0 | 1.0644 | 0.5783* | 0 | 1.0482 | 0.5777* |
| 1 | 3.3808 | 0.2280* | 1 | 2.7488 | 0.2250* |
| 2 | 1.7452 | 0.4920* | 0 | 2.5964 | 0.4292* |
| 0 | 4.8234 | 0.5519* | 0 | 4.4538 | 0.5475* |

**Table S4:**
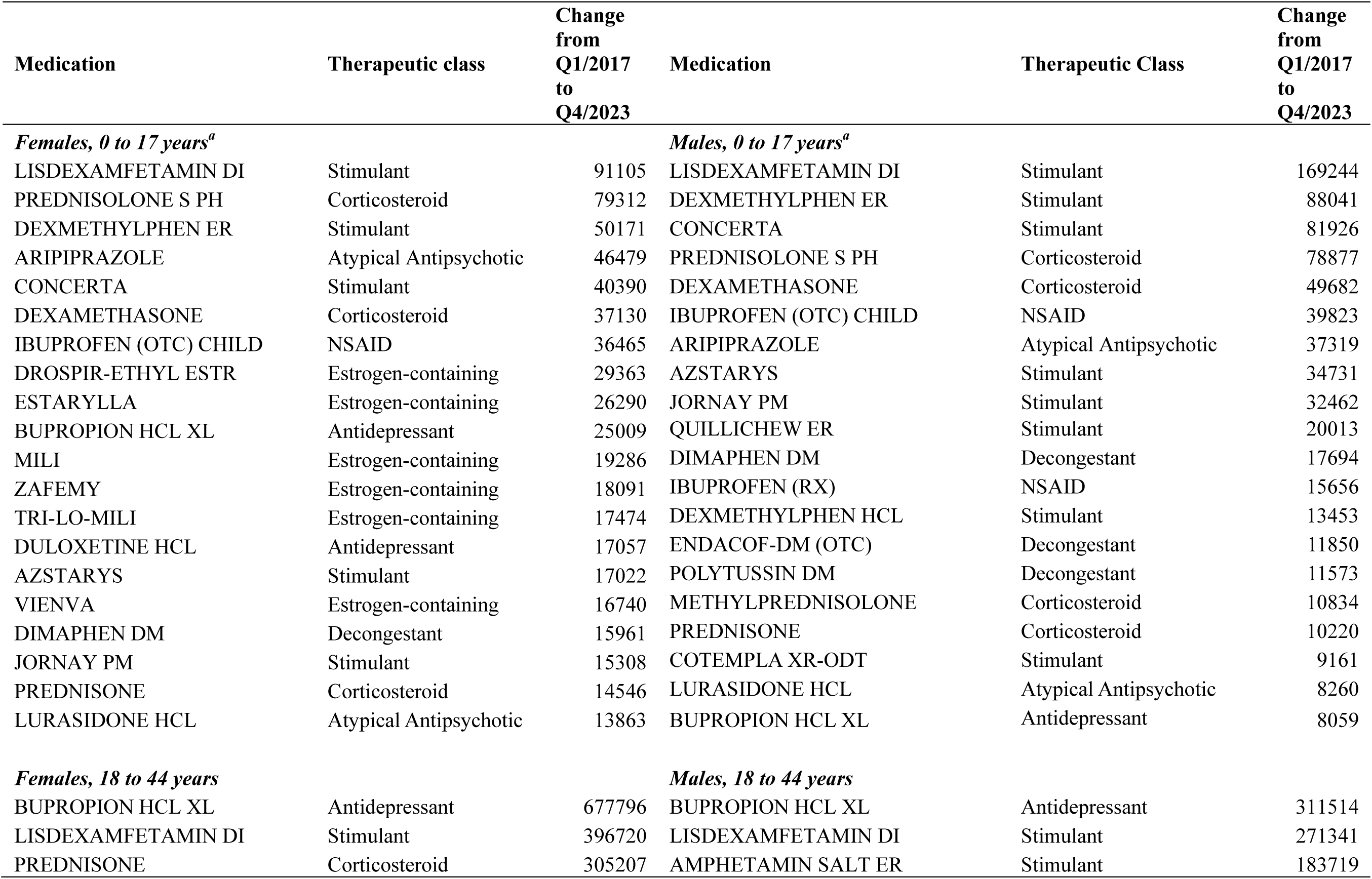
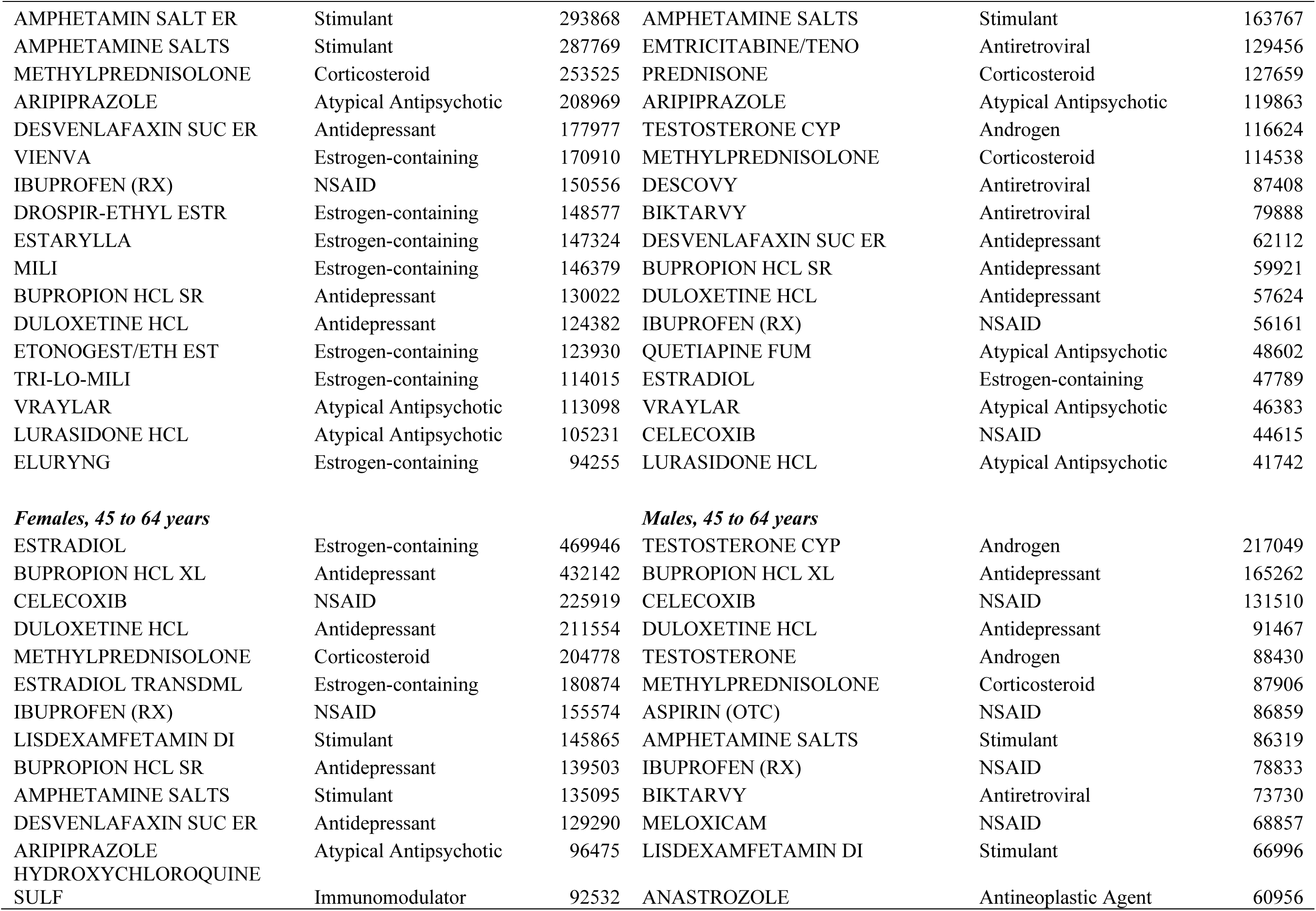
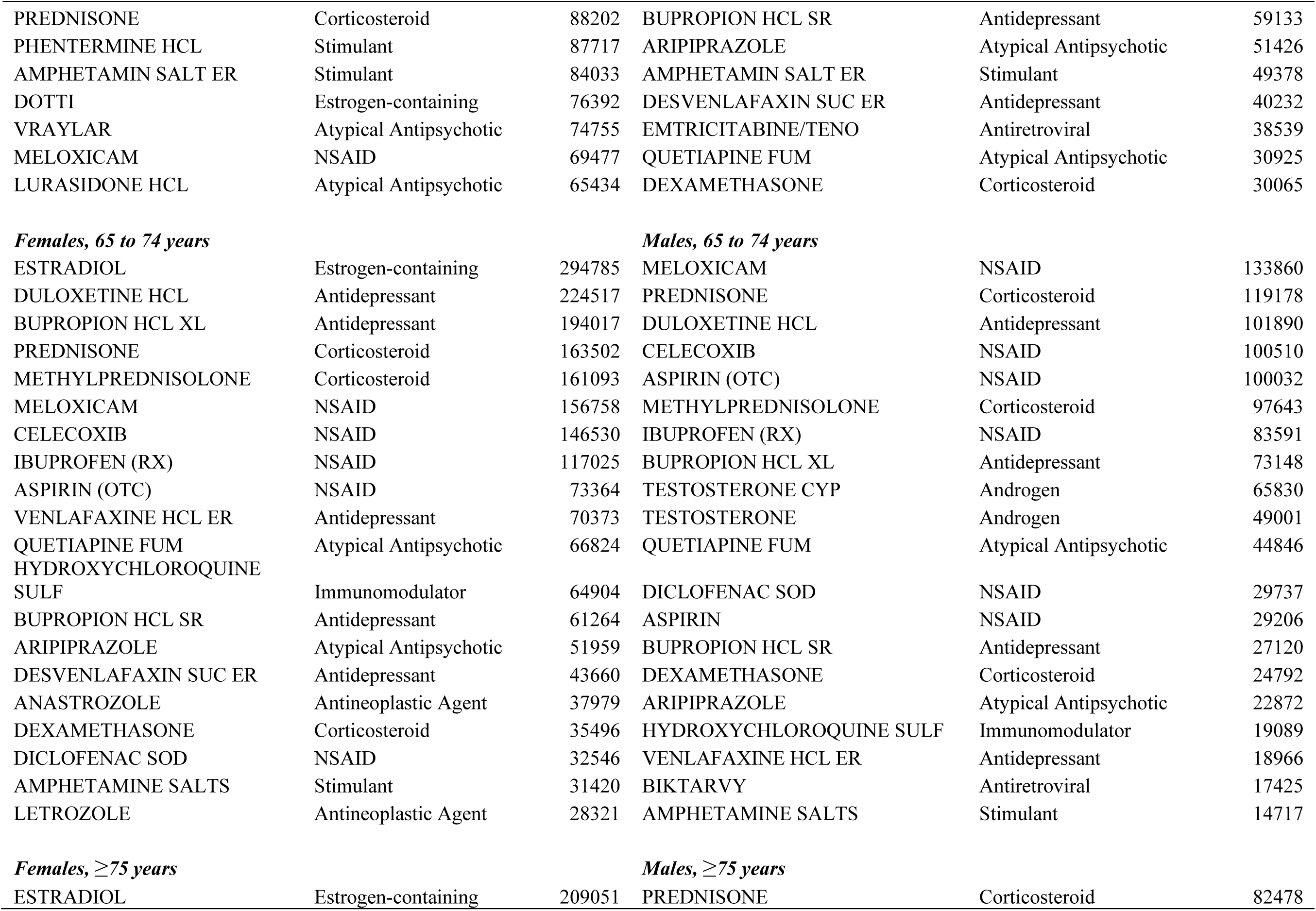
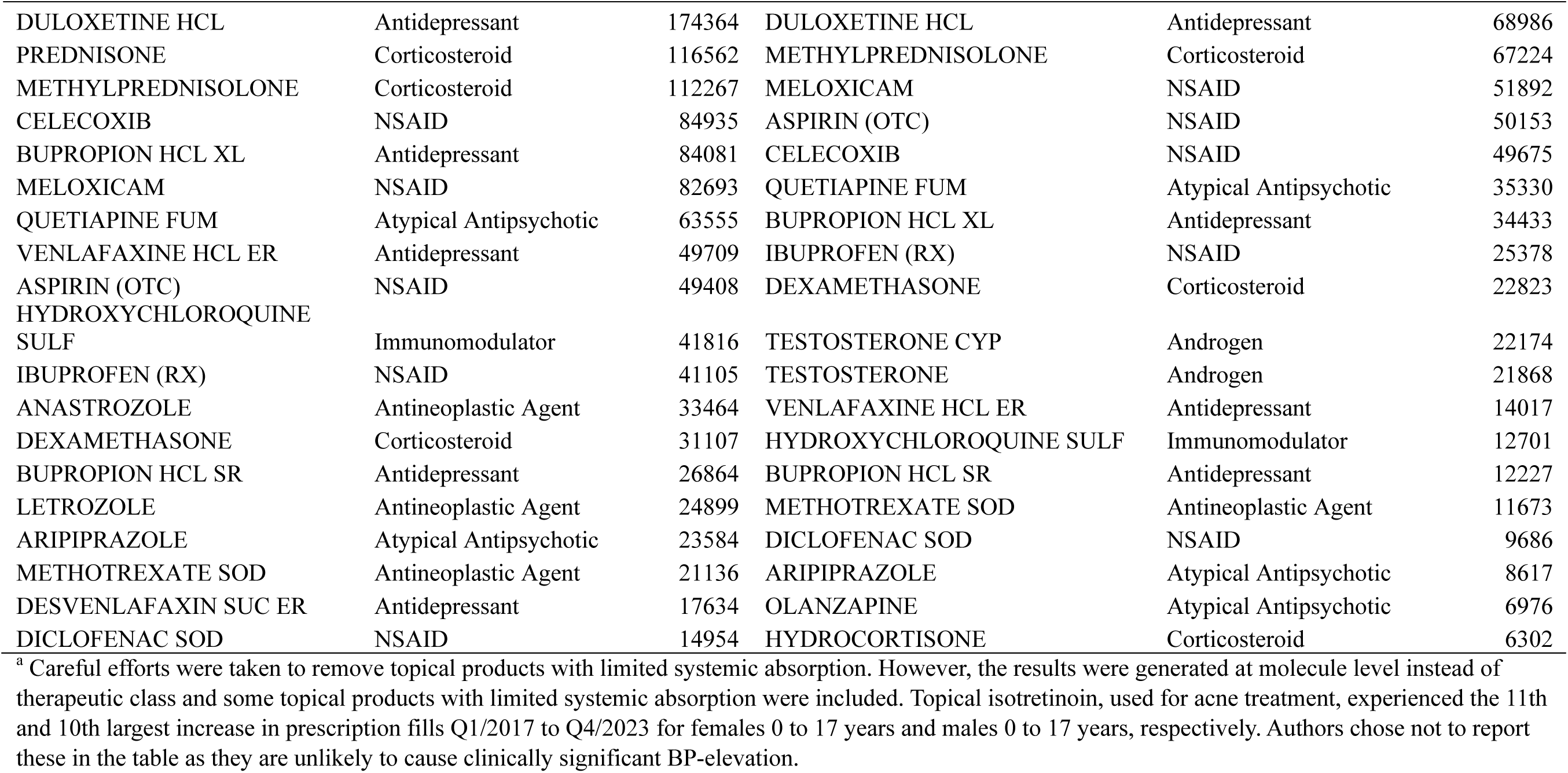
Medication and therapeutic class of the BP-elevating medications that experienced largest increase in prescription fills by sex and age group from Q1/2017 to Q4/2023, IQVIA Total Patient Tracker

**Table S5:**
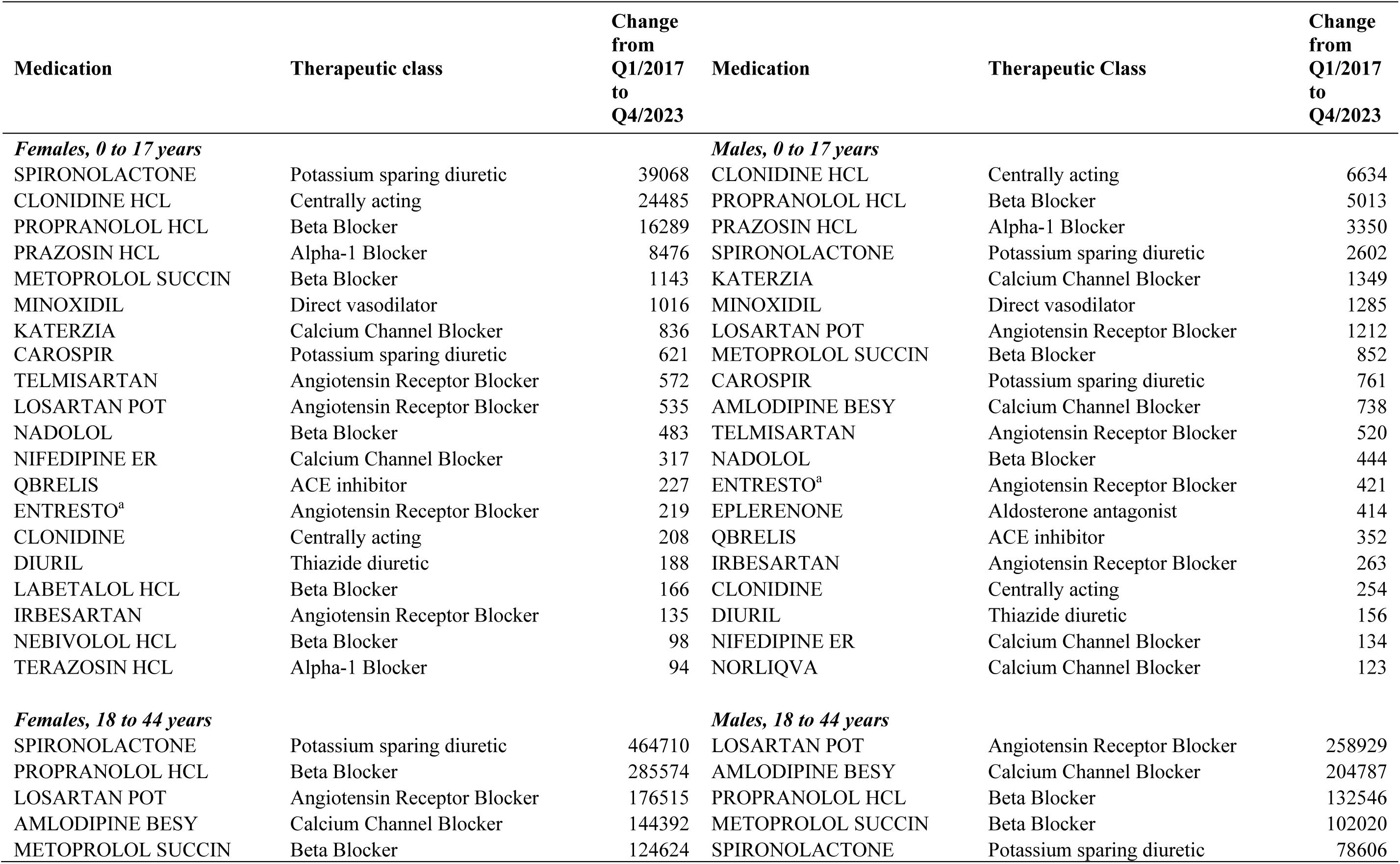
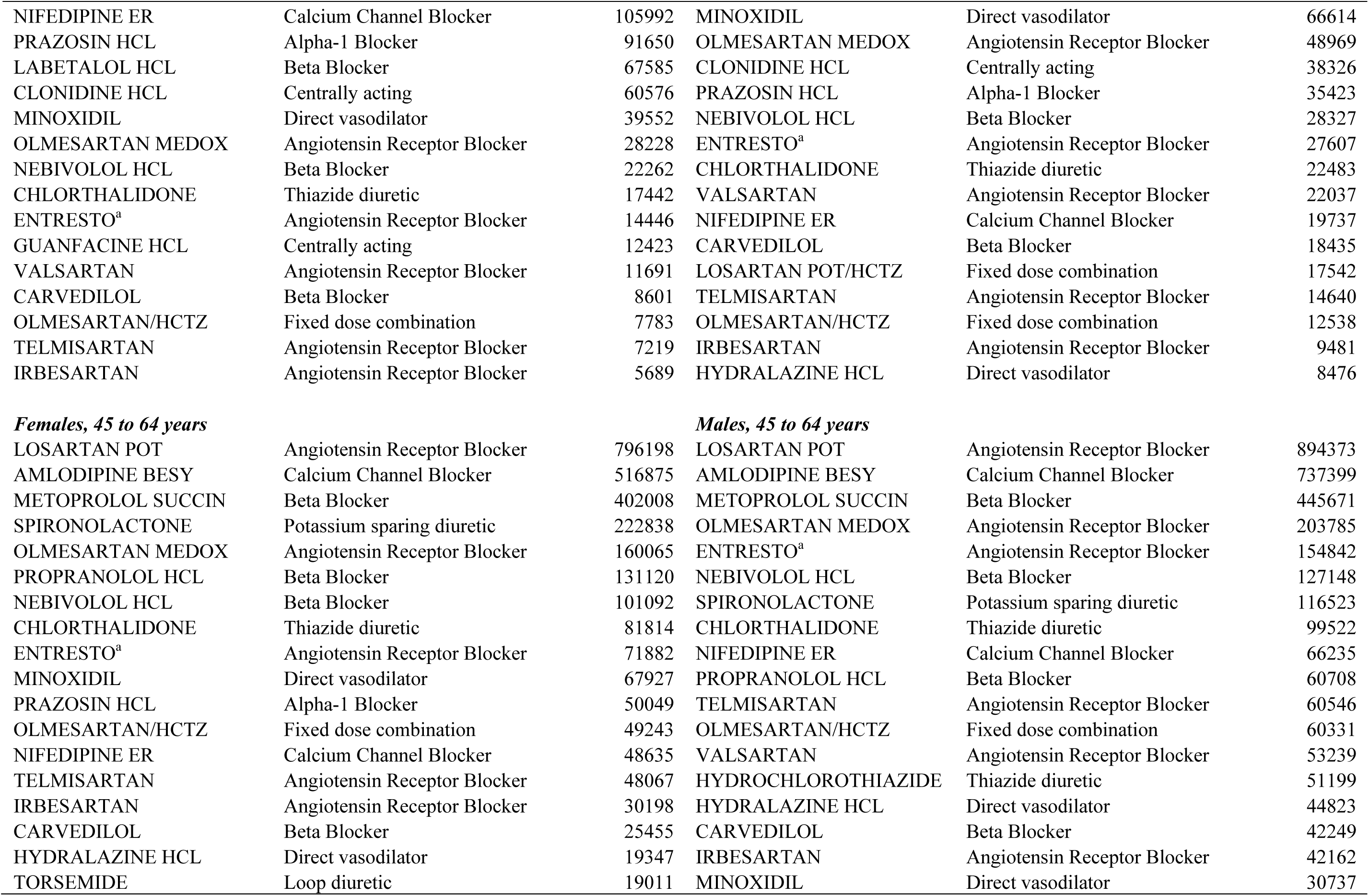
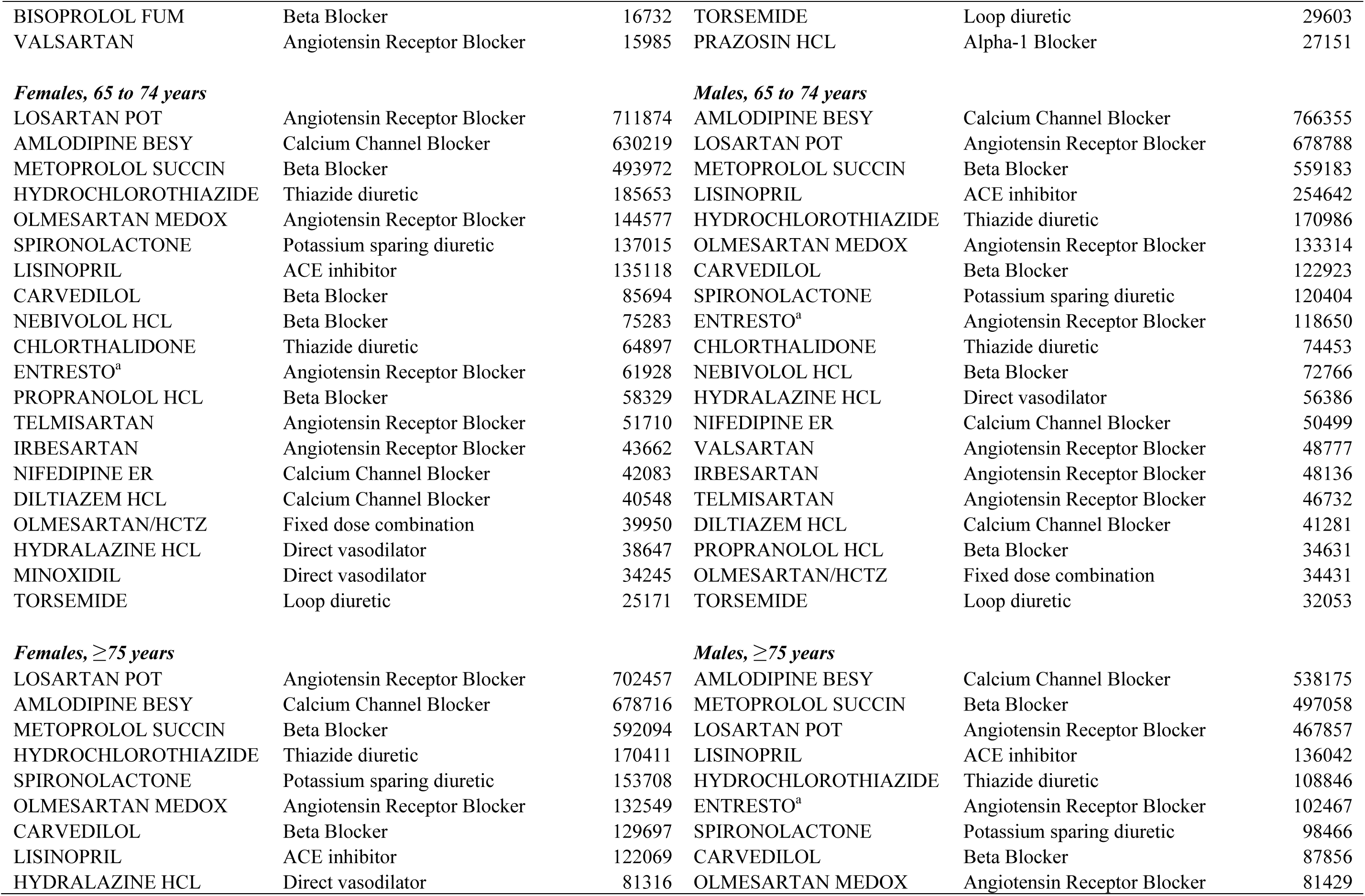
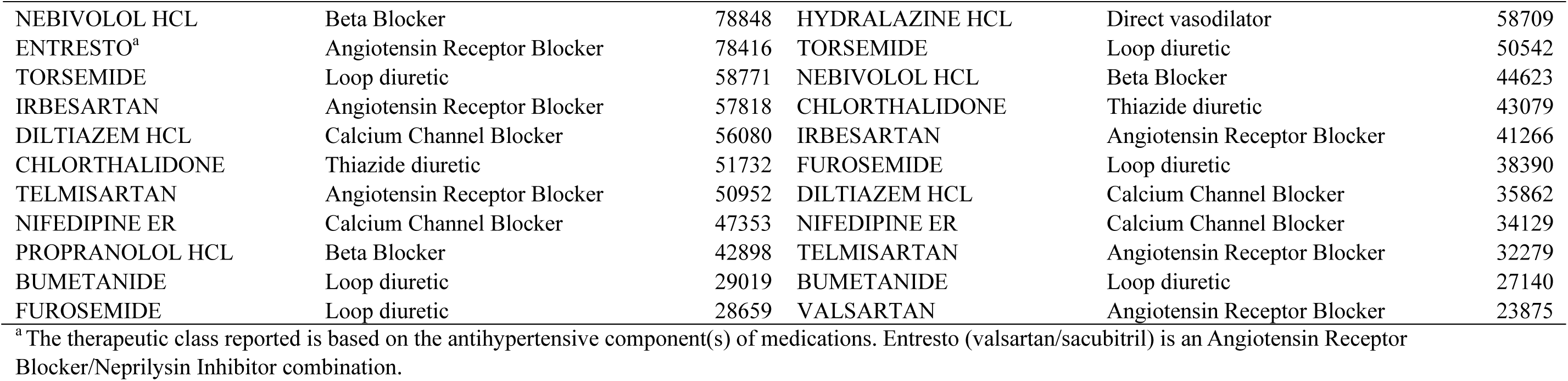
Medication and therapeutic class of the antihypertensive medications that experienced largest increase in prescription fills by sex and age group from Q1/2017 to Q4/2023, IQVIA Total Patient Tracker

| Medication | Therapeutic class | Change from Q1/2017 to Q4/2023 | Medication | Therapeutic Class | Change from Q1/2017 to Q4/2023 |
| --- | --- | --- | --- | --- | --- |
| *Females, 0 to 17 years* |  |  | *Males, 0 to 17 years* |  |  |
| SPIRONOLACTONE | Potassium sparing diuretic | 39068 | CLONIDINE HCL | Centrally acting | 6634 |
| CLONIDINE HCL | Centrally acting | 24485 | PROPRANOLOL HCL | Beta Blocker | 5013 |
| PROPRANOLOL HCL | Beta Blocker | 16289 | PRAZOSIN HCL | Alpha-1 Blocker | 3350 |
| PRAZOSIN HCL | Alpha-1 Blocker | 8476 | SPIRONOLACTONE | Potassium sparing diuretic | 2602 |
| METOPROLOL SUCCIN | Beta Blocker | 1143 | KATERZIA | Calcium Channel Blocker | 1349 |
| MINOXIDIL | Direct vasodilator | 1016 | MINOXIDIL | Direct vasodilator | 1285 |
| KATERZIA | Calcium Channel Blocker | 836 | LOSARTAN POT | Angiotensin Receptor Blocker | 1212 |
| CAROSPIR | Potassium sparing diuretic | 621 | METOPROLOL SUCCIN | Beta Blocker | 852 |
| TELMISARTAN | Angiotensin Receptor Blocker | 572 | CAROSPIR | Potassium sparing diuretic | 761 |
| LOSARTAN POT | Angiotensin Receptor Blocker | 535 | AMLODIPINE BESY | Calcium Channel Blocker | 738 |
| NADOLOL | Beta Blocker | 483 | TELMISARTAN | Angiotensin Receptor Blocker | 520 |
| NIFEDIPINE ER | Calcium Channel Blocker | 317 | NADOLOL | Beta Blocker | 444 |
| QBRELIS | ACE inhibitor | 227 | ENTRESTO<sup>a</sup> | Angiotensin Receptor Blocker | 421 |
| ENTRESTO<sup>a</sup> | Angiotensin Receptor Blocker | 219 | EPLERENONE | Aldosterone antagonist | 414 |
| CLONIDINE | Centrally acting | 208 | QBRELIS | ACE inhibitor | 352 |
| DIURIL | Thiazide diuretic | 188 | IRBESARTAN | Angiotensin Receptor Blocker | 263 |
| LABETALOL HCL | Beta Blocker | 166 | CLONIDINE | Centrally acting | 254 |
| IRBESARTAN | Angiotensin Receptor Blocker | 135 | DIURIL | Thiazide diuretic | 156 |
| NEBIVOLOL HCL | Beta Blocker | 98 | NIFEDIPINE ER | Calcium Channel Blocker | 134 |
| TERAZOSIN HCL | Alpha-1 Blocker | 94 | NORLIQVA | Calcium Channel Blocker | 123 |
| *Females, 18 to 44 years* |  |  | *Males, 18 to 44 years* |  |  |
| SPIRONOLACTONE | Potassium sparing diuretic | 464710 | LOSARTAN POT | Angiotensin Receptor Blocker | 258929 |
| PROPRANOLOL HCL | Beta Blocker | 285574 | AMLODIPINE BESY | Calcium Channel Blocker | 204787 |
| LOSARTAN POT | Angiotensin Receptor Blocker | 176515 | PROPRANOLOL HCL | Beta Blocker | 132546 |
| AMLODIPINE BESY | Calcium Channel Blocker | 144392 | METOPROLOL SUCCIN | Beta Blocker | 102020 |
| METOPROLOL SUCCIN | Beta Blocker | 124624 | SPIRONOLACTONE | Potassium sparing diuretic | 78606 |
| NIFEDIPINE ER | Calcium Channel Blocker | 105992 | MINOXIDIL | Direct vasodilator | 66614 |
| PRAZOSIN HCL | Alpha-1 Blocker | 91650 | OLMESARTAN MEDOX | Angiotensin Receptor Blocker | 48969 |
| LABETALOL HCL | Beta Blocker | 67585 | CLONIDINE HCL | Centrally acting | 38326 |
| CLONIDINE HCL | Centrally acting | 60576 | PRAZOSIN HCL | Alpha-1 Blocker | 35423 |
| MINOXIDIL | Direct vasodilator | 39552 | NEBIVOLOL HCL | Beta Blocker | 28327 |
| OLMESARTAN MEDOX | Angiotensin Receptor Blocker | 28228 | ENTRESTO<sup>a</sup> | Angiotensin Receptor Blocker | 27607 |
| NEBIVOLOL HCL | Beta Blocker | 22262 | CHLORTHALIDONE | Thiazide diuretic | 22483 |
| CHLORTHALIDONE | Thiazide diuretic | 17442 | VALSARTAN | Angiotensin Receptor Blocker | 22037 |
| ENTRESTO<sup>a</sup> | Angiotensin Receptor Blocker | 14446 | NIFEDIPINE ER | Calcium Channel Blocker | 19737 |
| GUANFACINE HCL | Centrally acting | 12423 | CARVEDILOL | Beta Blocker | 18435 |
| VALSARTAN | Angiotensin Receptor Blocker | 11691 | LOSARTAN POT/HCTZ | Fixed dose combination | 17542 |
| CARVEDILOL | Beta Blocker | 8601 | TELMISARTAN | Angiotensin Receptor Blocker | 14640 |
| OLMESARTAN/HCTZ | Fixed dose combination | 7783 | OLMESARTAN/HCTZ | Fixed dose combination | 12538 |
| TELMISARTAN | Angiotensin Receptor Blocker | 7219 | IRBESARTAN | Angiotensin Receptor Blocker | 9481 |
| IRBESARTAN | Angiotensin Receptor Blocker | 5689 | HYDRALAZINE HCL | Direct vasodilator | 8476 |
| ***Females, 45 to 64 years*** |  |  | ***Males, 45 to 64 years*** |  |  |
| LOSARTAN POT | Angiotensin Receptor Blocker | 796198 | LOSARTAN POT | Angiotensin Receptor Blocker | 894373 |
| AMLODIPINE BESY | Calcium Channel Blocker | 516875 | AMLODIPINE BESY | Calcium Channel Blocker | 737399 |
| METOPROLOL SUCCIN | Beta Blocker | 402008 | METOPROLOL SUCCIN | Beta Blocker | 445671 |
| SPIRONOLACTONE | Potassium sparing diuretic | 222838 | OLMESARTAN MEDOX | Angiotensin Receptor Blocker | 203785 |
| OLMESARTAN MEDOX | Angiotensin Receptor Blocker | 160065 | ENTRESTO<sup>a</sup> | Angiotensin Receptor Blocker | 154842 |
| PROPRANOLOL HCL | Beta Blocker | 131120 | NEBIVOLOL HCL | Beta Blocker | 127148 |
| NEBIVOLOL HCL | Beta Blocker | 101092 | SPIRONOLACTONE | Potassium sparing diuretic | 116523 |
| CHLORTHALIDONE | Thiazide diuretic | 81814 | CHLORTHALIDONE | Thiazide diuretic | 99522 |
| ENTRESTO<sup>a</sup> | Angiotensin Receptor Blocker | 71882 | NIFEDIPINE ER | Calcium Channel Blocker | 66235 |
| MINOXIDIL | Direct vasodilator | 67927 | PROPRANOLOL HCL | Beta Blocker | 60708 |
| PRAZOSIN HCL | Alpha-1 Blocker | 50049 | TELMISARTAN | Angiotensin Receptor Blocker | 60546 |
| OLMESARTAN/HCTZ | Fixed dose combination | 49243 | OLMESARTAN/HCTZ | Fixed dose combination | 60331 |
| NIFEDIPINE ER | Calcium Channel Blocker | 48635 | VALSARTAN | Angiotensin Receptor Blocker | 53239 |
| TELMISARTAN | Angiotensin Receptor Blocker | 48067 | HYDROCHLOROTHIAZIDE | Thiazide diuretic | 51199 |
| IRBESARTAN | Angiotensin Receptor Blocker | 30198 | HYDRALAZINE HCL | Direct vasodilator | 44823 |
| CARVEDILOL | Beta Blocker | 25455 | CARVEDILOL | Beta Blocker | 42249 |
| HYDRALAZINE HCL | Direct vasodilator | 19347 | IRBESARTAN | Angiotensin Receptor Blocker | 42162 |
| TORSEMIDE | Loop diuretic | 19011 | MINOXIDIL | Direct vasodilator | 30737 |
| BISOPROLOL FUM | Beta Blocker | 16732 | TORSEMIDE | Loop diuretic | 29603 |
| VALSARTAN | Angiotensin Receptor Blocker | 15985 | PRAZOSIN HCL | Alpha-1 Blocker | 27151 |
| *Females, 65 to 74 years* |  |  | *Males, 65 to 74 years* |  |  |
| LOSARTAN POT | Angiotensin Receptor Blocker | 711874 | AMLODIPINE BESY | Calcium Channel Blocker | 766355 |
| AMLODIPINE BESY | Calcium Channel Blocker | 630219 | LOSARTAN POT | Angiotensin Receptor Blocker | 678788 |
| METOPROLOL SUCCIN | Beta Blocker | 493972 | METOPROLOL SUCCIN | Beta Blocker | 559183 |
| HYDROCHLOROTHIAZIDE | Thiazide diuretic | 185653 | LISINOPRIL | ACE inhibitor | 254642 |
| OLMESARTAN MEDOX | Angiotensin Receptor Blocker | 144577 | HYDROCHLOROTHIAZIDE | Thiazide diuretic | 170986 |
| SPIRONOLACTONE | Potassium sparing diuretic | 137015 | OLMESARTAN MEDOX | Angiotensin Receptor Blocker | 133314 |
| LISINOPRIL | ACE inhibitor | 135118 | CARVEDILOL | Beta Blocker | 122923 |
| CARVEDILOL | Beta Blocker | 85694 | SPIRONOLACTONE | Potassium sparing diuretic | 120404 |
| NEBIVOLOL HCL | Beta Blocker | 75283 | ENTRESTO<sup>a</sup> | Angiotensin Receptor Blocker | 118650 |
| CHLORTHALIDONE | Thiazide diuretic | 64897 | CHLORTHALIDONE | Thiazide diuretic | 74453 |
| ENTRESTO<sup>a</sup> | Angiotensin Receptor Blocker | 61928 | NEBIVOLOL HCL | Beta Blocker | 72766 |
| PROPRANOLOL HCL | Beta Blocker | 58329 | HYDRALAZINE HCL | Direct vasodilator | 56386 |
| TELMISARTAN | Angiotensin Receptor Blocker | 51710 | NIFEDIPINE ER | Calcium Channel Blocker | 50499 |
| IRBESARTAN | Angiotensin Receptor Blocker | 43662 | VALSARTAN | Angiotensin Receptor Blocker | 48777 |
| NIFEDIPINE ER | Calcium Channel Blocker | 42083 | IRBESARTAN | Angiotensin Receptor Blocker | 48136 |
| DILTIAZEM HCL | Calcium Channel Blocker | 40548 | TELMISARTAN | Angiotensin Receptor Blocker | 46732 |
| OLMESARTAN/HCTZ | Fixed dose combination | 39950 | DILTIAZEM HCL | Calcium Channel Blocker | 41281 |
| HYDRALAZINE HCL | Direct vasodilator | 38647 | PROPRANOLOL HCL | Beta Blocker | 34631 |
| MINOXIDIL | Direct vasodilator | 34245 | OLMESARTAN/HCTZ | Fixed dose combination | 34431 |
| TORSEMIDE | Loop diuretic | 25171 | TORSEMIDE | Loop diuretic | 32053 |
| *Females, ≥75 years* |  |  | *Males, ≥75 years* |  |  |
| LOSARTAN POT | Angiotensin Receptor Blocker | 702457 | AMLODIPINE BESY | Calcium Channel Blocker | 538175 |
| AMLODIPINE BESY | Calcium Channel Blocker | 678716 | METOPROLOL SUCCIN | Beta Blocker | 497058 |
| METOPROLOL SUCCIN | Beta Blocker | 592094 | LOSARTAN POT | Angiotensin Receptor Blocker | 467857 |
| HYDROCHLOROTHIAZIDE | Thiazide diuretic | 170411 | LISINOPRIL | ACE inhibitor | 136042 |
| SPIRONOLACTONE | Potassium sparing diuretic | 153708 | HYDROCHLOROTHIAZIDE | Thiazide diuretic | 108846 |
| OLMESARTAN MEDOX | Angiotensin Receptor Blocker | 132549 | ENTRESTO<sup>a</sup> | Angiotensin Receptor Blocker | 102467 |
| CARVEDILOL | Beta Blocker | 129697 | SPIRONOLACTONE | Potassium sparing diuretic | 98466 |
| LISINOPRIL | ACE inhibitor | 122069 | CARVEDILOL | Beta Blocker | 87856 |
| HYDRALAZINE HCL | Direct vasodilator | 81316 | OLMESARTAN MEDOX | Angiotensin Receptor Blocker | 81429 |
| NEBIVOLOL HCL | Beta Blocker | 78848 | HYDRALAZINE HCL | Direct vasodilator | 58709 |
| ENTRESTO<sup>a</sup> | Angiotensin Receptor Blocker | 78416 | TORSEMIDE | Loop diuretic | 50542 |
| TORSEMIDE | Loop diuretic | 58771 | NEBIVOLOL HCL | Beta Blocker | 44623 |
| IRBESARTAN | Angiotensin Receptor Blocker | 57818 | CHLORTHALIDONE | Thiazide diuretic | 43079 |
| DILTIAZEM HCL | Calcium Channel Blocker | 56080 | IRBESARTAN | Angiotensin Receptor Blocker | 41266 |
| CHLORTHALIDONE | Thiazide diuretic | 51732 | FUROSEMIDE | Loop diuretic | 38390 |
| TELMISARTAN | Angiotensin Receptor Blocker | 50952 | DILTIAZEM HCL | Calcium Channel Blocker | 35862 |
| NIFEDIPINE ER | Calcium Channel Blocker | 47353 | NIFEDIPINE ER | Calcium Channel Blocker | 34129 |
| PROPRANOLOL HCL | Beta Blocker | 42898 | TELMISARTAN | Angiotensin Receptor Blocker | 32279 |
| BUMETANIDE | Loop diuretic | 29019 | BUMETANIDE | Loop diuretic | 27140 |
| FUROSEMIDE | Loop diuretic | 28659 | VALSARTAN | Angiotensin Receptor Blocker | 23875 |

<sup>a</sup> The therapeutic class reported is based on the antihypertensive component(s) of medications. Entresto (valsartan/sacubitril) is an Angiotensin Receptor Blocker/Neprilysin Inhibitor combination.

**Table S6:**
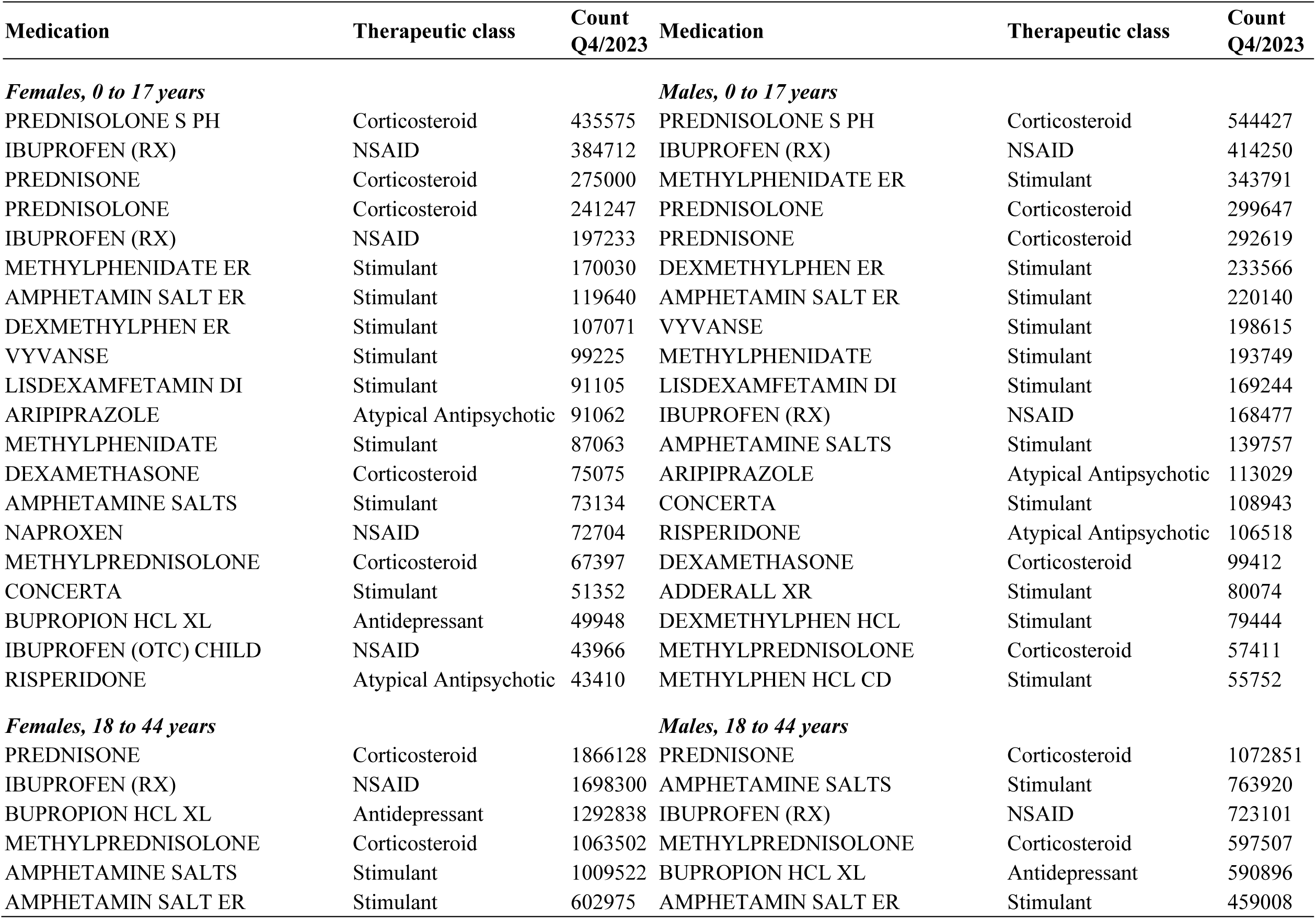
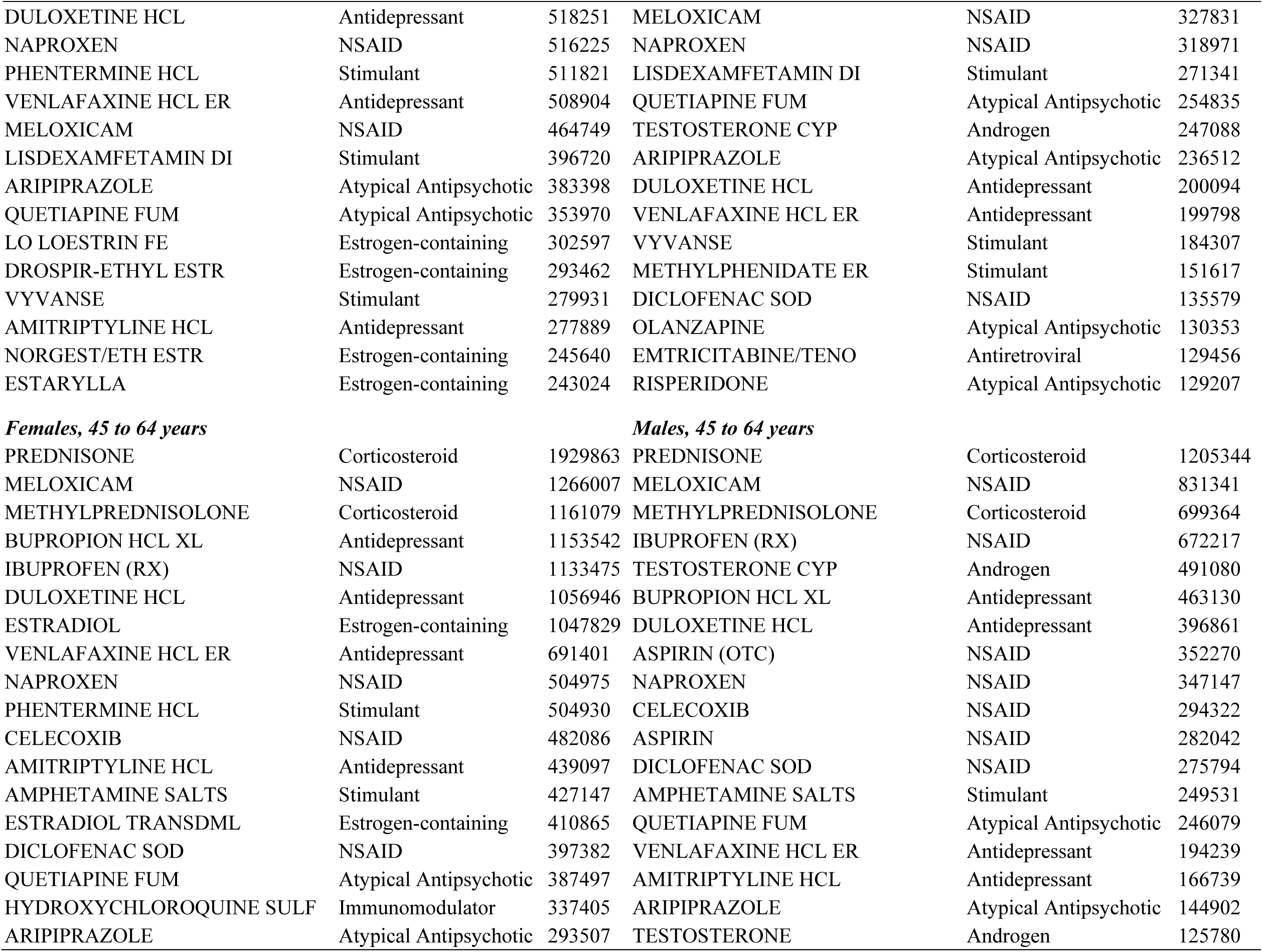
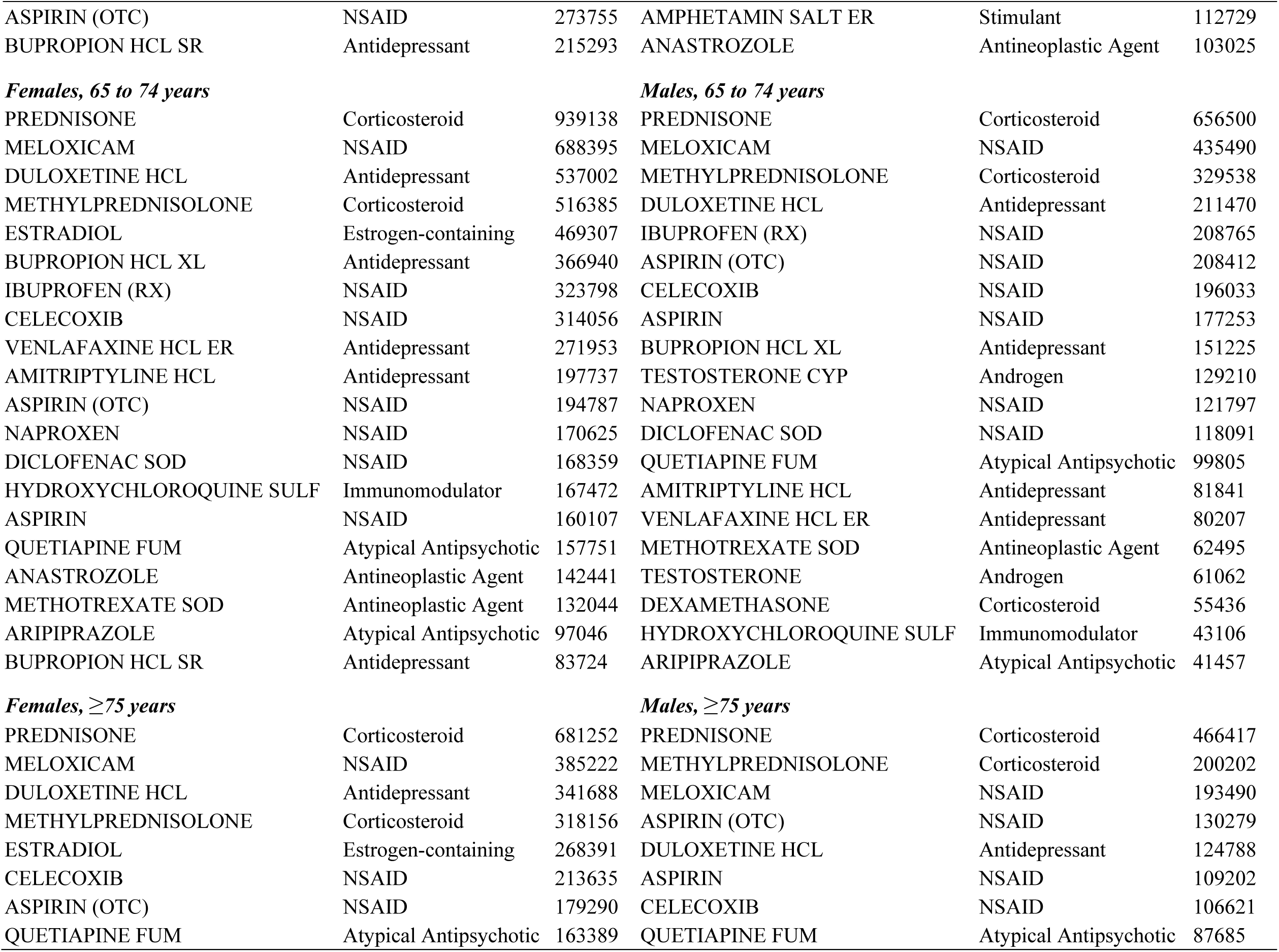
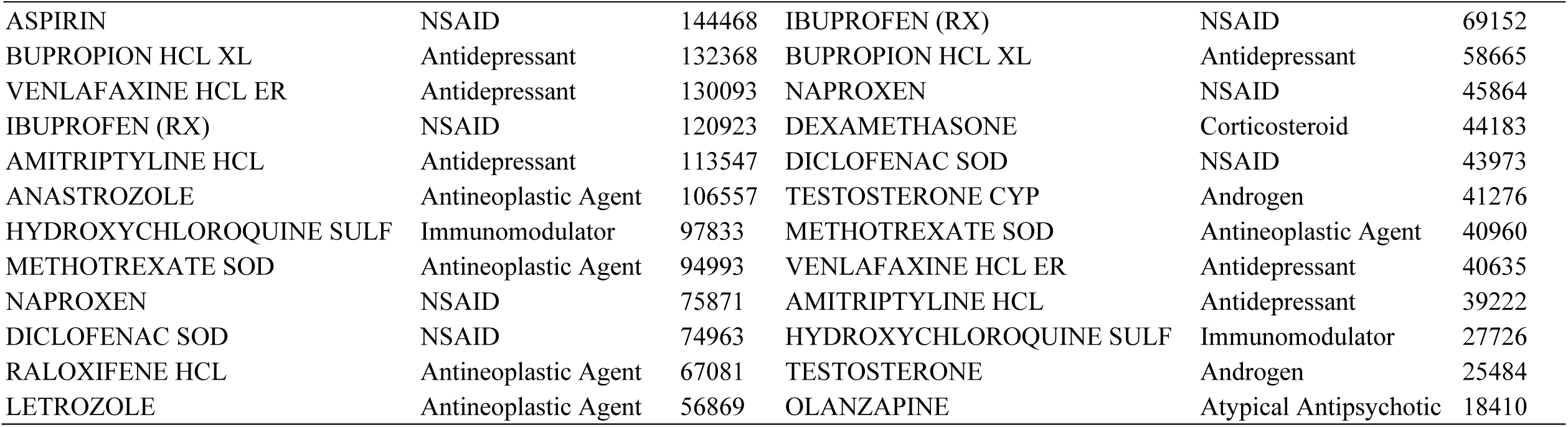
Medication and therapeutic class of the BP-elevating medications with highest number of prescription fills by sex and age group in Q4/2023, IQVIA Total Patient Tracker

**Table S7:** Medication and therapeutic class of the antihypertensive medications with highest number of prescription fills by sex and age group in Q4/2023, IQVIA Total Patient Tracker

| Medication | Therapeutic class | Count<br>Q4/2023 | Medication | Therapeutic class | Count<br>Q4/2023 |
| --- | --- | --- | --- | --- | --- |
| ***Females, 0 to 17 years*** |  |  | ***Males, 0 to 17 years*** |  |  |
| CLONIDINE HCL | Centrally acting | 147194 | CLONIDINE HCL | Centrally acting | 275839 |
| SPIRONOLACTONE | Potassium sparing diuretic | 55451 | GUANFACINE HCL | Centrally acting | 130000 |
| GUANFACINE HCL | Centrally acting | 52015 | PROPRANOLOL HCL | Beta Blocker | 19067 |
| PROPRANOLOL HCL | Beta Blocker | 36909 | LISINOPRIL | ACE inhibitor | 15945 |
| PRAZOSIN HCL | Alpha-1 Blocker | 17552 | ENALAPRIL MAL | ACE inhibitor | 9584 |
| LISINOPRIL | ACE inhibitor | 8882 | AMLODIPINE BESY | Calcium Channel Blocker | 8739 |
| ENALAPRIL MAL | ACE inhibitor | 7140 | PRAZOSIN HCL | Alpha-1 Blocker | 7245 |
| AMLODIPINE BESY | Calcium Channel Blocker | 6449 | FUROSEMIDE | Loop diuretic | 6916 |
| ATENOLOL | Beta Blocker | 6326 | SPIRONOLACTONE | Loop diuretic | 6145 |
| FUROSEMIDE | Loop diuretic | 5871 | ATENOLOL | Beta Blocker | 5300 |
| METOPROLOL SUCCIN | Beta Blocker | 3021 | LOSARTAN POT | Angiotensin Receptor Blocker | 4809 |
| LOSARTAN POT | Angiotensin Receptor Blocker | 2845 | METOPROLOL SUCCIN | Beta Blocker | 2539 |
| NADOLOL | Beta Blocker | 2331 | NADOLOL | Beta Blocker | 2246 |
| HYDROCHLOROTHIAZIDE | Thiazide diuretic | 1283 | HYDROCHLOROTHIAZIDE | Thiazide diuretic | 1726 |
| METOPROLOL TART | Beta Blocker | 1123 | MINOXIDIL | Direct vasodilator | 1354 |
| MINOXIDIL | Direct vasodilator | 1048 | KATERZIA | Calcium Channel Blocker | 1349 |
| BENAZEPRIL HCL | ACE inhibitor | 935 | CARVEDILOL | Beta Blocker | 1292 |
| NIFEDIPINE ER | Calcium Channel Blocker | 881 | CLONIDINE | Centrally acting | 1181 |
| KATERZIA | Calcium Channel Blocker | 836 | BENAZEPRIL HCL | ACE inhibitor | 1054 |
| DIURIL | Thiazide diuretic | 769 | DIURIL | Thiazide diuretic | 942 |
| ***Females, 18 to 44 years*** |  |  | ***Males, 18 to 44 years*** |  |  |
| SPIRONOLACTONE | Potassium sparing diuretic | 872348 | LISINOPRIL | ACE inhibitor | 831620 |
| PROPRANOLOL HCL | Beta Blocker | 546239 | AMLODIPINE BESY | Calcium Channel Blocker | 633869 |
| AMLODIPINE BESY | Calcium Channel Blocker | 484185 | LOSARTAN POT | Angiotensin Receptor Blocker | 495211 |
| LISINOPRIL | ACE inhibitor | 473787 | METOPROLOL SUCCIN | Beta Blocker | 282295 |
| LOSARTAN POT | Angiotensin Receptor Blocker | 356734 | PROPRANOLOL HCL | Beta Blocker | 267743 |
| HYDROCHLOROTHIAZIDE | Thiazide diuretic | 339435 | HYDROCHLOROTHIAZIDE | Thiazide diuretic | 239617 |
| METOPROLOL SUCCIN | Beta Blocker | 329825 | CLONIDINE HCL | Centrally acting | 174697 |
| LABETALOL HCL | Beta Blocker | 191723 | LISINOPRIL/HCTZ | Fixed dose combination | 151539 |
| CLONIDINE HCL | Centrally acting | 182962 | SPIRONOLACTONE | Potassium sparing diuretic | 131895 |
| PRAZOSIN HCL | Alpha-1 Blocker | 169297 | CARVEDILOL | Beta Blocker | 119047 |
| NIFEDIPINE ER | Calcium Channel Blocker | 166151 | LOSARTAN POT/HCTZ | Fixed dose combination | 108888 |
| METOPROLOL TART | Beta Blocker | 122684 | METOPROLOL TART | Beta Blocker | 102880 |
| FUROSEMIDE | Loop diuretic | 117964 | FUROSEMIDE | Loop diuretic | 77349 |
| LISINOPRIL/HCTZ | Fixed dose combination | 99819 | MINOXIDIL | Direct vasodilator | 71504 |
| LOSARTAN POT/HCTZ | Fixed dose combination | 85916 | PRAZOSIN HCL | Alpha-1 Blocker | 68796 |
| ATENOLOL | Beta Blocker | 80602 | ATENOLOL | Beta Blocker | 61418 |
| CARVEDILOL | Beta Blocker | 79709 | VALSARTAN | Angiotensin Receptor Blocker | 61109 |
| CHLORTHALIDONE | Thiazide diuretic | 46790 | OLMESARTAN MEDOX | Angiotensin Receptor Blocker | 58393 |
| MINOXIDIL | Direct vasodilator | 40900 | CHLORTHALIDONE | Thiazide diuretic | 51159 |
| VALSARTAN | Angiotensin Receptor Blocker | 36166 | NIFEDIPINE ER | Calcium Channel Blocker | 41906 |
| ***Females, 45 to 64 years*** |  |  | ***Males, 45 to 64 years*** |  |  |
| AMLODIPINE BESY | Calcium Channel Blocker | 2373740 | LISINOPRIL | ACE inhibitor | 3110829 |
| LISINOPRIL | ACE inhibitor | 2252851 | AMLODIPINE BESY | Calcium Channel Blocker | 2921428 |
| LOSARTAN POT | Angiotensin Receptor Blocker | 1992193 | LOSARTAN POT | Angiotensin Receptor Blocker | 2050941 |
| HYDROCHLOROTHIAZIDE | Thiazide diuretic | 1571096 | METOPROLOL SUCCIN | Beta Blocker | 1440407 |
| METOPROLOL SUCCIN | Beta Blocker | 1317697 | HYDROCHLOROTHIAZIDE | Thiazide diuretic | 1112971 |
| FUROSEMIDE | Loop diuretic | 609657 | CARVEDILOL | Beta Blocker | 723904 |
| SPIRONOLACTONE | Potassium sparing diuretic | 600942 | LISINOPRIL/HCTZ | Fixed dose combination | 660509 |
| LISINOPRIL/HCTZ | Fixed dose combination | 578842 | METOPROLOL TART | Beta Blocker | 600966 |
| METOPROLOL TART | Beta Blocker | 551531 | FUROSEMIDE | Loop diuretic | 513197 |
| LOSARTAN POT/HCTZ | Fixed dose combination | 490902 | LOSARTAN POT/HCTZ | Fixed dose combination | 473500 |
| CARVEDILOL | Beta Blocker | 472397 | SPIRONOLACTONE | Potassium sparing diuretic | 349508 |
| PROPRANOLOL HCL | Beta Blocker | 428901 | VALSARTAN | Angiotensin Receptor Blocker | 287140 |
| ATENOLOL | Beta Blocker | 349616 | OLMESARTAN MEDOX | Angiotensin Receptor Blocker | 261095 |
| TRIAMTERENE/HCTZ | Fixed dose combination | 249982 | ATENOLOL | Beta Blocker | 258732 |
| VALSARTAN | Angiotensin Receptor Blocker | 225777 | CHLORTHALIDONE | Thiazide diuretic | 230906 |
| CLONIDINE HCL | Centrally acting | 223830 | PROPRANOLOL HCL | Beta Blocker | 215643 |
| CHLORTHALIDONE | Thiazide diuretic | 210965 | HYDRALAZINE HCL | Direct vasodilator | 193884 |
| OLMESARTAN MEDOX | Angiotensin Receptor Blocker | 206563 | NIFEDIPINE ER | Calcium Channel Blocker | 185629 |
| NIFEDIPINE ER | Calcium Channel Blocker | 159090 | CLONIDINE HCL | Centrally acting | 182150 |
| DILTIAZEM HCL | Calcium Channel Blocker | 156467 | AMLODIP BES/BENAZ HCL | Fixed dose combination | 181842 |
| **Females, 65 to 74 years** |  |  | **Males, 65 to 74 years** |  |  |
| AMLODIPINE BESY | Calcium Channel Blocker | 2042210 | AMLODIPINE BESY | Calcium Channel Blocker | 2047614 |
| LOSARTAN POT | Angiotensin Receptor Blocker | 1644259 | LISINOPRIL | ACE inhibitor | 1828706 |
| LISINOPRIL | ACE inhibitor | 1560881 | LOSARTAN POT | Angiotensin Receptor Blocker | 1381379 |
| METOPROLOL SUCCIN | Beta Blocker | 1212471 | METOPROLOL SUCCIN | Beta Blocker | 1257325 |
| HYDROCHLOROTHIAZIDE | Thiazide diuretic | 1094635 | HYDROCHLOROTHIAZIDE | Thiazide diuretic | 748562 |
| FUROSEMIDE | Loop diuretic | 597298 | CARVEDILOL | Beta Blocker | 617925 |
| METOPROLOL TART | Beta Blocker | 533629 | METOPROLOL TART | Beta Blocker | 556908 |
| CARVEDILOL | Beta Blocker | 484265 | FUROSEMIDE | Loop diuretic | 516223 |
| LISINOPRIL/HCTZ | Fixed dose combination | 347702 | LISINOPRIL/HCTZ | Fixed dose combination | 340471 |
| LOSARTAN POT/HCTZ | Fixed dose combination | 332947 | SPIRONOLACTONE | Potassium sparing diuretic | 270614 |
| SPIRONOLACTONE | Potassium sparing diuretic | 328881 | LOSARTAN POT/HCTZ | Fixed dose combination | 259846 |
| ATENOLOL | Beta Blocker | 328393 | ATENOLOL | Beta Blocker | 213155 |
| VALSARTAN | Angiotensin Receptor Blocker | 207627 | VALSARTAN | Angiotensin Receptor Blocker | 199638 |
| TRIAMTERENE/HCTZ | Fixed dose combination | 203198 | OLMESARTAN MEDOX | Angiotensin Receptor Blocker | 163034 |
| DILTIAZEM HCL | Calcium Channel Blocker | 189023 | HYDRALAZINE HCL | Direct vasodilator | 162156 |
| OLMESARTAN MEDOX | Angiotensin Receptor Blocker | 179950 | DILTIAZEM HCL | Calcium Channel Blocker | 153704 |
| PROPRANOLOL HCL | Beta Blocker | 178346 | CHLORTHALIDONE | Thiazide diuretic | 145105 |
| HYDRALAZINE HCL | Direct vasodilator | 149955 | ENTRESTO<sup>a</sup> | Angiotensin Receptor Blocker | 130561 |
| CHLORTHALIDONE | Thiazide diuretic | 146081 | NIFEDIPINE ER | Calcium Channel Blocker | 130015 |
| NIFEDIPINE ER | Calcium Channel Blocker | 133191 | AMLODIP BES/BENAZ HCL | Fixed dose combination | 116515 |
| **Females, ≥75 years** |  |  | **Males, ≥75 years** |  |  |
| AMLODIPINE BESY | Calcium Channel Blocker | 2234939 | AMLODIPINE BESY | Calcium Channel Blocker | 1449233 |
| LOSARTAN POT | Angiotensin Receptor Blocker | 1591330 | LISINOPRIL | ACE inhibitor | 1076944 |
| METOPROLOL SUCCIN | Beta Blocker | 1404260 | METOPROLOL SUCCIN | Beta Blocker | 1060242 |
| LISINOPRIL | ACE inhibitor | 1269216 | LOSARTAN POT | Angiotensin Receptor Blocker | 953516 |
| FUROSEMIDE | Loop diuretic | 988134 | FUROSEMIDE | Loop diuretic | 682525 |
| HYDROCHLOROTHIAZIDE | Thiazide diuretic | 854629 | CARVEDILOL | Beta Blocker | 504714 |
| METOPROLOL TART | Beta Blocker | 668455 | METOPROLOL TART | Beta Blocker | 480491 |
| CARVEDILOL | Beta Blocker | 596682 | HYDROCHLOROTHIAZIDE | Thiazide diuretic | 468836 |
| SPIRONOLACTONE | Potassium sparing diuretic | 349881 | SPIRONOLACTONE | Potassium sparing diuretic | 223277 |
| ATENOLOL | Beta Blocker | 340267 | ATENOLOL | Beta Blocker | 168322 |
| DILTIAZEM HCL | Calcium Channel Blocker | 281519 | HYDRALAZINE HCL | Direct vasodilator | 155522 |
| HYDRALAZINE HCL | Direct vasodilator | 245929 | LISINOPRIL/HCTZ | Fixed dose combination | 148794 |
| LOSARTAN POT/HCTZ | Fixed dose combination | 230679 | DILTIAZEM HCL | Calcium Channel Blocker | 139858 |
| VALSARTAN | Angiotensin Receptor Blocker | 214331 | VALSARTAN | Angiotensin Receptor Blocker | 135762 |
| LISINOPRIL/HCTZ | Fixed dose combination | 200940 | DOXAZOSIN MESY | Alpha-1 Blocker | 130022 |
| OLMESARTAN MEDOX | Angiotensin Receptor Blocker | 172875 | LOSARTAN POT/HCTZ | Fixed dose combination | 128480 |
| TRIAMTERENE/HCTZ | Fixed dose combination | 170304 | ENTRESTO<sup>a</sup> | Angiotensin Receptor Blocker | 112533 |
| NIFEDIPINE ER | Calcium Channel Blocker | 160854 | OLMESARTAN MEDOX | Angiotensin Receptor Blocker | 100728 |
| CLONIDINE HCL | Centrally acting | 131841 | NIFEDIPINE ER | Calcium Channel Blocker | 97200 |
| PROPRANOLOL HCL | Beta Blocker | 130104 | TORSEMIDE | Loop diuretic | 97184 |

<sup>a</sup>The therapeutic class reported is based on the antihypertensive component(s) of medications. Entresto (valsartan/sacubitril) is an Angiotensin Receptor Blocker/Neprilysin Inhibitor combination.

